# A gene network implicated in the joint-muscle pain, brain fog, chronic fatigue, and bowel irregularity of Ehlers-Danlos and “long” COVID19 syndromes

**DOI:** 10.1101/2023.03.24.23287706

**Authors:** Golder N. Wilson

## Abstract

**Objectives:** Characterization of tissue laxity *<u>and</u>* dysautonomia symptoms in Ehlers-Danlos syndrome (EDS) uncovered similarities with those of post-infectious SARS-CoV-2 or long COVID19, prompting detailed comparison of their findings and influencing genes.

**Methods:** Holistic assessment of 1261 EDS outpatients for 120 history-physical findings populated a deidentified database that includes 568 patients with 317 variant genes obtained by commercial NextGen sequencing. Findings were compared to 15 of long COVID19 compiled in an extensive review, genes to 104 associated with COVID19 severity in multiple molecular studies.

**Results:** Fifteen symptoms common to Ehlers-Danlos versus long COVID19 ranged from brain fog (27-80 versus 30-70%), chronic fatigue (38-91; 30-60%), dyspnea (32-52; 29-52%) to irritable bowel (67-89; 14-78%), muscle weakness (22-49; 15-25%), and arthritis (32-94; 15-27%). Genes relevant to EDS included 6 identical to those influencing COVID19 severity (*F2*, *LIFR*, *NLRP3*, *STAT1*, *T1CAM1*, *TNFRSF13B*) and 18 similar including *POLG*-*POLD4*, *SLC6A2-SLC6A20*, and *NFKB1-NFKB2*. Both gene sets had broad genomic distribution, many mitochondrial genes influencing EDS and many involved with immunity-inflammation modifying COVID19 severity. Recurring DNA variants in EDS that merit evaluation in COVID19 resistance include those impacting connective tissue elements--51 in *COL5* (joint), 29 in *COL1/2/9/11* (bone), 13 in *COL3* (vessel), and 18 in *FBN1* (vessel-heart)--or neural function--93 in mitochondrial DNA, 28 in *COL6/12*, 16 in *SCN9A/10A/11A*, 14 in *POLG*, and 11 in genes associated with porphyria.

**Conclusions:** Holistic ascertainment of finding pattern and exome variation in EDS defined tissue laxity, neuromuscular, and autonomic correlations that transcend single abnormalities or types. Implied networks of nuclear and mitochondrial genes are linked to findings like brain fog, fatigue, and frailty in EDS, their similarity to long COVID19 supporting shared therapies for disorders affecting a minimum 0.1% of the global population.

## Introduction

Study of the human being, limited by causal foibles of chance and necessity, can nevertheless take advantage of a large organism privileged by centuries of detailed observation. Human systems biology can begin with the Review of Systems required for medical evaluation, a holistic approach nicely complemented by NextGen detailing of genome sequence change [1–3]. While the contingencies of disease pattern will never match the controlled insights from experimental study, holistic documentation of symptoms and their translation into pathogenetic mechanisms can focus molecular investigation. Such is the case when the full panoply of tissue laxity [4–8], autonomic [9–11], and neuromuscular [12–13] findings are ascertained in connective tissue dysplasias [8], appreciation of Ehlers-Danlos *<u>syndrome</u>* (EDS) linking its genetic variation to central articulo-autonomic dysplasia mechanisms [11] instead of peripheral phenotypes [4–8].

Although initial analyses of connective tissue disorders focused on tethering proteins like collagens [1–7], its necessary role in protist to metazoan transitions [14] requires that connecting tissue be medium [15–21] and message [12, 22–28]. The result in complex organisms that combine internal homeostasis with external movement is a diversity of constraining and connecting structures [skin, joint, skeleton—15-21] permeated by wired [nerve 12-13, 22-25], moving [muscle 26-28] and circulating [heart-vessel 7] parts. All of these tissues including many blood components have a common developmental origin [mesoderm, mesenchyme--29] that reflects an evolutionary drive for cell connection.

The inevitable consequence of this integrated anatomy is a reciprocal relationship between the systems that constrain/contain body or blood [20,21] and the nervous system that coordinates their functions [22,23]. Disposition to tissue laxity will not only cause wear-and-tear osteoarthritis and skeletal bends from gravity [deformations like scoliosis, 4-8] but also will provoke adrenergic response to restore cerebral circulation deprived by vessel distensibility and lower body blood pooling [9–12]. Repeated adrenergic stimulation, evident even in those with minimal or benign joint hypermobility [9], produces the brain fog, stress response, and chronic fatigue of postural orthostatic tachycardia syndrome [30–32], the reactive allergic [32–33], immune [23,34], and inflammatory [35] symptoms of mast cell activation [32–33], and, through cholinergic suppression, the irregularity, reflux, and swallowing difficulties of irritable bowel syndrome [36].

These aspects of dysautonomia are receiving renewed emphasis after being recognized in patients recovering from SARS-CoV-2 [37–43], an RNA beta-coronavirus producing a respiratory disease syndrome called coronavirus disease 2019 or COVID19 [44–46]. The virus has caused over 650 million infections and 6.6 million deaths worldwide since its emergence from China in late 2019 [45], 6.2% of infections associated with autonomic and respiratory symptoms that persist for weeks or months after the initial illness has improved [43]. These post-infectious symptoms have become known as a post-acute COVID19 sequelae (PACS) or long COVID19 syndrome that can occur after a 1 to 2-week course of mild, severe, or asymptomatic disease [37,38]. The frequency and timing of long COVID19 symptoms, like those of EDS, are highly variable as shown by the 59 of 303 studies qualifying for review by Deer et al. [37].

Individual variability and heritability of certain self-reported COVID19 symptoms [47] coupled with descriptions of X-linked COVID19 susceptibility [48] prompted studies to define genes modulating COVID19 susceptibility [48–50]. Recently reviewed [50] are top-down approaches analyzing interactive gene modules [51] or molecular pathways [52–54] altered by COVID 19 infection and bottom-up studies focusing on individual genes using whole genome association, DNA sequencing, and CRISPR ablation analyses. Overlap of symptoms between EDS [9–11], acute [55–59], and post-acute COVID19 [37–43] suggested that their contributing genes might be similar, prompting analysis that could foster application of proven therapies [6–11] to a novel and globally escalating disorder.

## Methods

### Patients

Patients were evaluated in a private medical genetics practice after whole exome sequencing became economically feasible via preliminary ascertainment of insurance coverage by the GeneDx© Company. EDS and developmental disability patients were seen from July of 2011 to August of 2017, those with EDS being the sole practice focus from the latter date through October of 2020. Clinical evaluations and DNA testing of EDS patients were performed as described preliminarily [2,11]; the 1656 diagnosed with EDS expanded to 1899, the 710 with systematic evaluations for the 120 findings in Supporting Information S1 Table to 1261, the 727 with DNA testing to 967 (article Table 1). The most recent 243 EDS patients, including 153 with DNA testing and 90 (59%) with positive results, were evaluated by telemedicine/online interaction after the private office closed in July 2018. Patients with obvious diagnoses of Marfan, Loeys-Dietz, or skeletal dysplasias were excluded. Patients with developmental disability and/or autism had different evaluations in the private office as previously described [2]. Part-time appointment at Texas Tech University Health Sciences Centers included separate genetics clinic and laboratory administrative work at that Center while coordinating the Dallas private practice.

**Table 1.** Ehlers-Danlos syndrome patients and their DNA testing.

| Patients group | All EDS | Female | Male | F>10.5y | F<10.5y | DNA all | DNA+ | DNA- | No EDS |
| --- | --- | --- | --- | --- | --- | --- | --- | --- | --- |
| **Total number (No.) T** | **1899** | **1553** | **346** | **1446** | **107** | **967** | **573** | **394** | **80** |
| **No. systematic eval (%T)** | **1261 (66)** | **1064 (69)** | **197 (57)** | **1020 (71)** | **44 (41)** | **854 (88)** | **568 (99)** | **286 (73)** | **64 (80)** |
| **No. routine eval (%T)** | **638 (34)** | **489 (31)** | **149( 43)** | **426 (29)** | **63 (59)** | **113 (12)** | **5 (1.0)** | **108 (27)** | **16 (20)** |
| **Age of eval (years X±SD)** | **29±14** | **30±13** | **21±13<sup>a</sup>** | **31±13** | **6.2±2.8<sup>b</sup>** | **29±14** | **29±14** | **30±13** | **16±9.1<sup>c</sup>** |
| **Age severe (years X±SD)** | **17±9.2** | **17±9.0** | **14±9.9<sup>a</sup>** | **18±8.8** | **4.8±2.3<sup>b</sup>** | **17±9.7** | **16±9.7** | **17±9.6** | **14±7.6<sup>c</sup>** |
| **Average Hx of 80 (X±SD)** | **34±10** | **36±9.8** | **26±8.0<sup>a</sup>** | **37±9.4** | **22±9.4<sup>b</sup>** | **35±9.9** | **35±9.9** | **35±10** | **7.2±1.3<sup>c</sup>** |
| **Average PE of 40 (X±SD)** | **18±4.7** | **19±4.5** | **17±4.8<sup>a</sup>** | **19±4.5** | **16±3.1<sup>b</sup>** | **19±4.8** | **18±4.7** | **19±4.8** | **7.6±1.3<sup>c</sup>** |

### DNA testing

Patients and/or families were given forms to consent for medical genetic evaluation/treatment and anonymous sharing of DNA results from whole exome sequencing (WES) during patient intake, counseled regarding ambiguous, incomplete, or incidental/secondary findings [60] and consented to send their insurance information to the GeneDx© Company for estimates of out-of-pocket costs. GeneDx genetic counselors obtained out-of-pocket cost estimates for testing, completed requisitions with generic consents for de-identified data and secondary finding sharing, and coordinated cheek swab sampling of patient and parents when available. Results using standard methods for whole exome sequencing [61,62] with independent [63] or conjoint [64] microarray analysis were obtained by fax and/or internet portal. Results were provided with counsel by the author at follow-up clinic visits.

### Patient and DNA databases

The 1979 EDS and 725 developmental disability patients having outpatient evaluations were entered into a password-protected MS Excel© GW patient database as approved by the North Texas IRB (centered at Medical City Hospital, Dallas) in 2014 (exempt protocol number 2014-054). Data on 305 EDS patients seen before 2014 were entered after approval, 68 entered as dictated by protocol guidelines after its closure on 19-12-2018 when the author closed the Medical City office). The 1261 EDS patients with systematic evaluations were transferred to a more comprehensive EDS database with history-physical findings, specification of those related, sex, age range (2.5 years under age 10, 10 years for those over age 10.1 years), type of visit (online or clinic), referral (self, specialist, or primary physician), and DNA results. The specific DNA variants found in 568 EDS patients with positive testing results were extracted and listed in S3 Table of Supplementary Information, the rest of the information available (with indication of positive/negative but not specific DNA results) as an EDS1261GW1-23 database by request to the author. Those interested in further research can match clinical and DNA findings through concordant patient numbers in S3 Table and the requested database, accessing EDS patients who are often anxious to participate in validating research. DNA variants in 82 patients with developmental disability were separately extracted from the larger database and listed in S4 Table to allow comparison with the DNA variation in EDS patients.

Genes modifying COVID19 infection were taken from articles obtained by PubMed searches using those terms conducted through December 2022. Genes and article references were entered into the parallel Excel database as listed in the S5 Table of Supporting information, their previously associated diseases taken from OMIM as described next.

### Classification of gene products, impacts on tissue elements/processes

The Online Mendelian Inheritance in Man (OMIM) at www.omim.org, accessed from June 2021 to January 2023, provides reference (M) numbers for genes variant in EDS or developmental disability patients and for those related to COVID19 severity (respective S2 and S4-S5 Tables of the Supporting Information). Diseases associated with each gene are also referenced by (M) numbers, condensed lists of symptoms provided for the EDS- [S2 Table] and COVID19-related [S5 Table] but not for the disability-related diseases because the latter are less-relevant conditions with developmental-intellectual disability and/or autistic behaviors.

Each gene product is classified by function (e. g., enzyme, receptor, membrane channel as shown in the legend to Fig. 3) based on its description in the OMIM entry. Classification of genes by impact on tissue element or process per the Fig. 2 and 4, S2 Table legends relies on symptoms of their associated diseases in S2 and S4 Tables, assignments more arbitrary since many associated diseases affect multiple systems and many genes are associated with more than one disease (M+ symbol in S2 and S4-S5 Tables).

**Fig 1.**
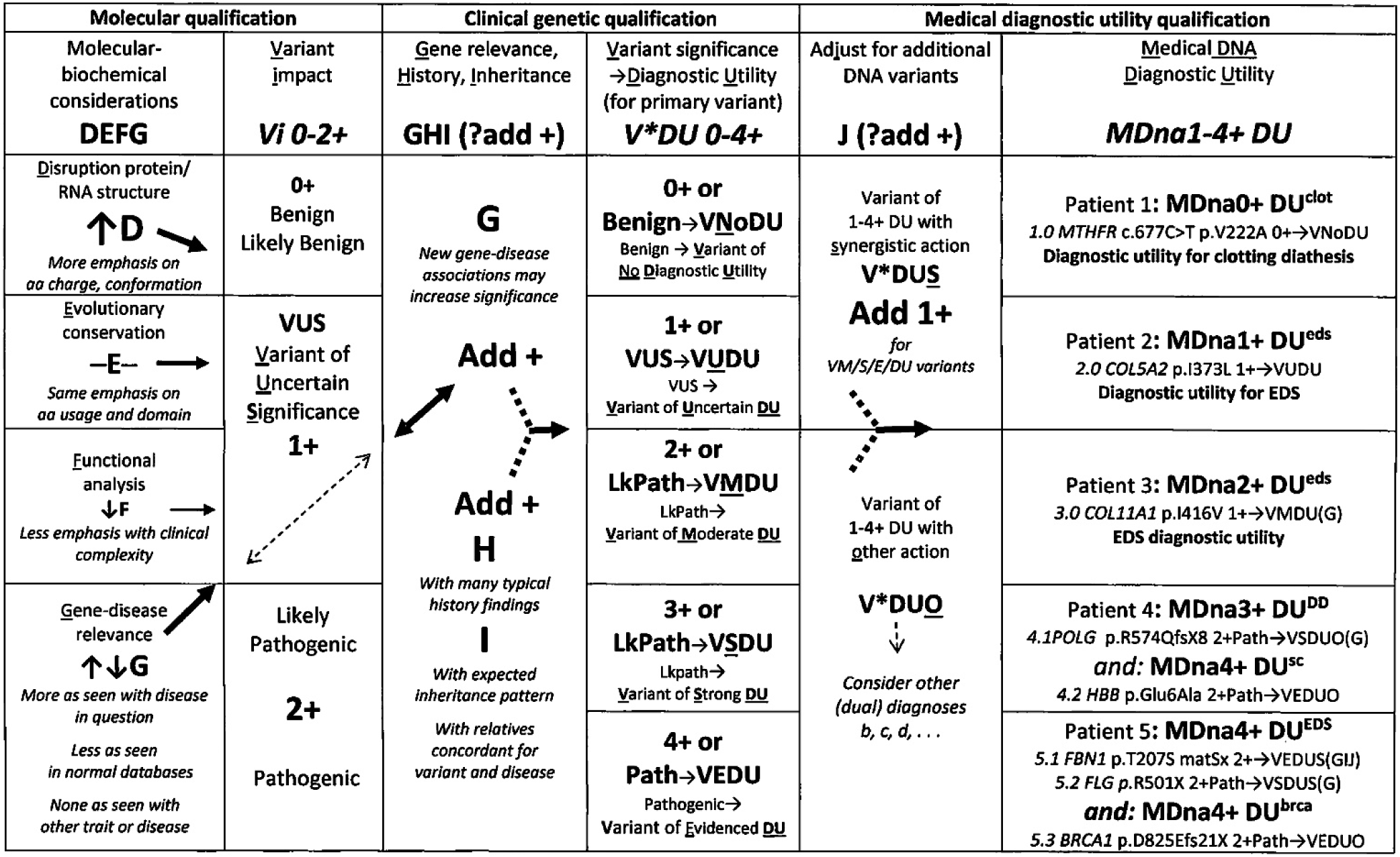
Clinical protocol for DNA variant qualification. Clinical DNA variant (column 4) and 1-4+ medical diagnostic utilities (last column) are added to consensus qualifications (column 2) as discussed in the text, DNA/protein change and gene abbreviations except for *MTHFR* (methylene tetrahydrofolate reductase) and *HBB* (beta-globin) are explained in S2, S3 Tables.

**Fig 2.**
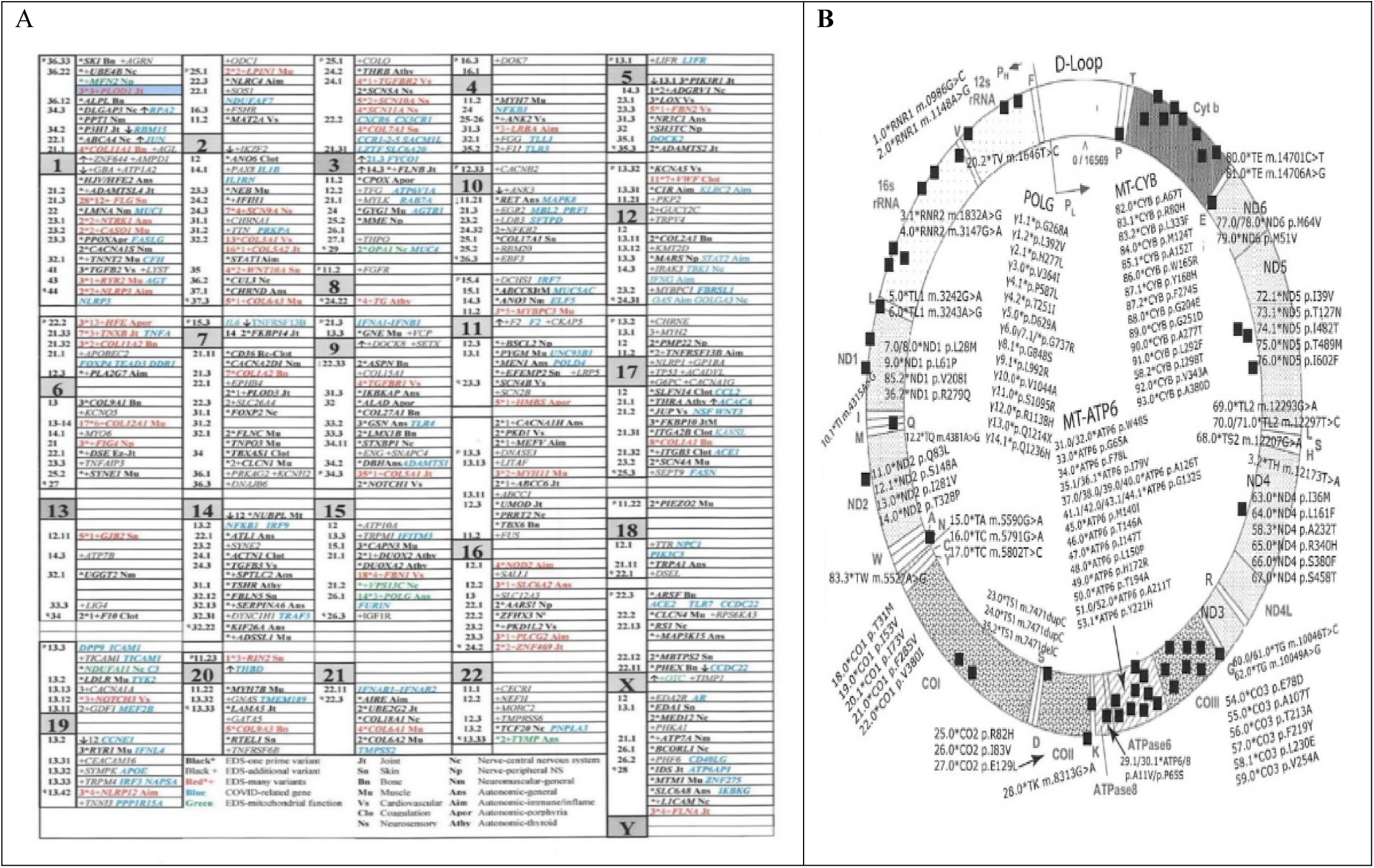
Mapping of nuclear and mitochondrial genes associated with EDS and COVID19 severity. A. Nuclear genes from S2 Table are shown with numbers of primary variants in bold followed by *, of additional variants in italics followed by +, recurring variants in red, genes encoding products transported to mitochondria in green, genes related to COVID19 severity in blue (S5 Table); gene abbreviations, exact loci in S2 and S6 Tables, chromosome sizes modified for display by factors ≅ x1/2 for numbers 4-5-9, x1/4 for 8, x2/3 for 10; x1.1 for 14-21-X; x 1.3 for 22, x 1.7 for 20; x2 for 16-17-19 [63]; B, Primary DNA variants are described by DNA (m.) or protein (p.) position, additional ones by 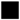 --see variant details in S2 Table for Fig 2A, S3 Table adds 473 to variant numbers in Fig. 2B, patient 473 is number 1 above; map from MITOMAP [89].

**Fig 3.**
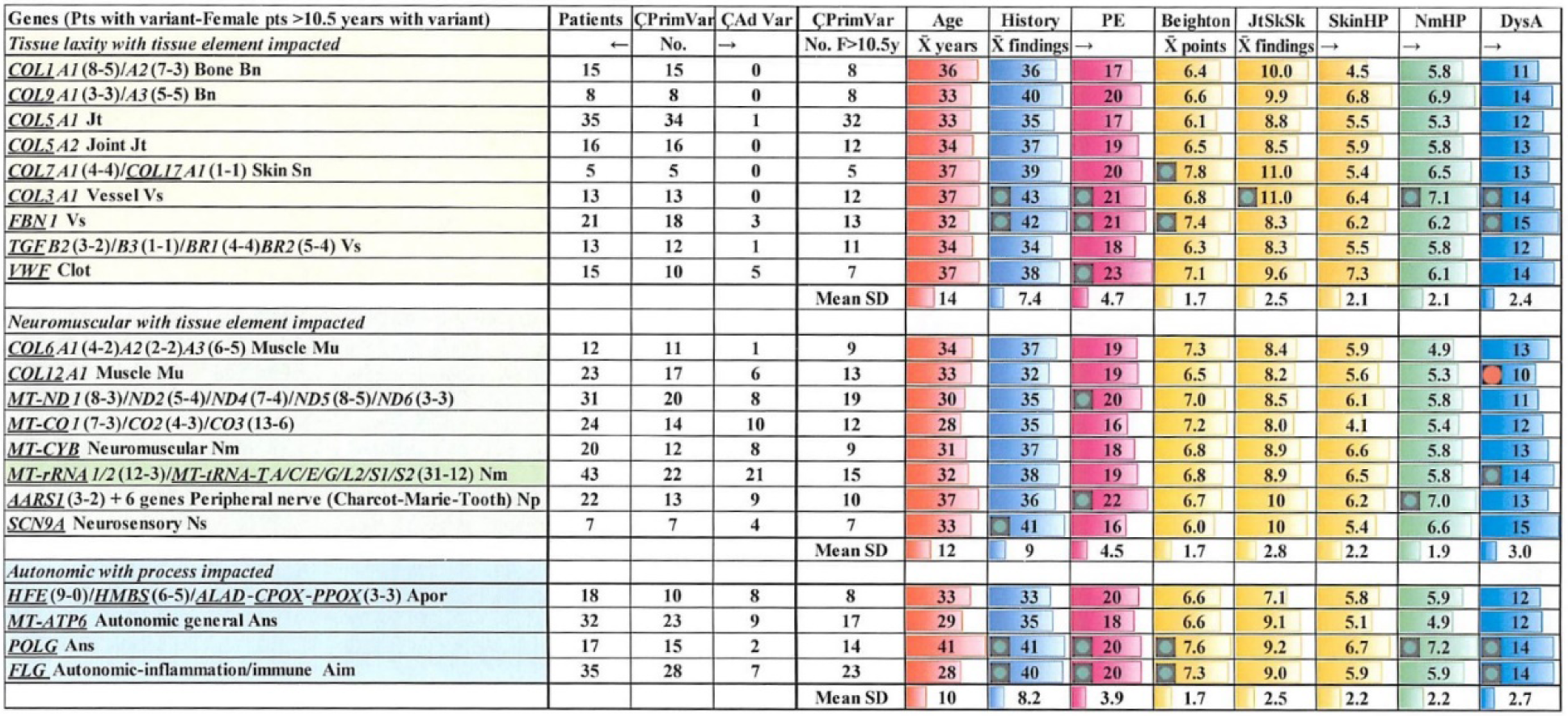
Similar EDS-dysautonomia finding numbers in patients with recurring gene variants. Gene abbreviations and patient numbers are from S2 Table, the first four columns showing all patients with variants, then with primary variants (PrimVar), with only additional variants (AdVar), and female patients over age 10.5 years with variants—only the latter qualify for finding comparison; finding categories from S1 Table include mean age (years), history (of 80), physical (of 40), Beighton (of 9), joint-skeletal by history-by physical (JtSkSk of 21), skin (of 11), neuromuscular by history-physical (NmHP of 16), and dysautonomia (DysA of 20) 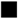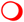, significant difference p<0.05, see Methods; Ç, with; X^-^, mean; SD, standard deviation.

**Fig. 4.**
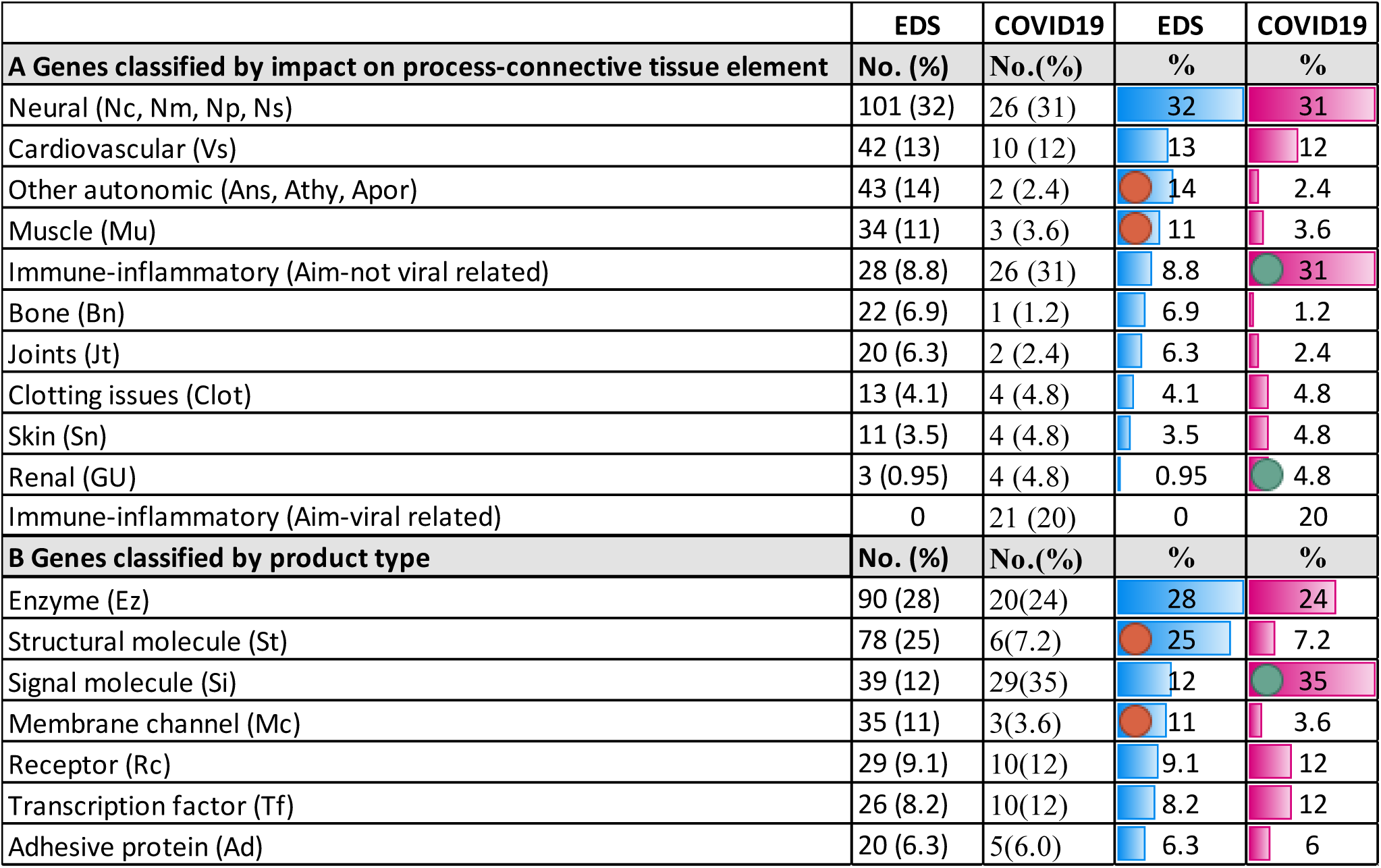
Genes relevant to EDS or COVID19 infection by tissue element or product type. **A**, process/connective tissue element relations (box, Fig 2A bottom) are from associated diseases (S2, S5 Tables); COVID19 percentages of 83 genes after 21 impacting viral-related processes were subtracted; **B**, gene product functions are explained in the legend to Table S2, COVID19 percentages are of all 104 genes listed in Table S4 (the *PNPLA3* gene associated with gastrointestinal disease is not listed); significantly (p < 0.05) lower 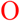 or higher 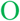 proportions (see Methods).

### Statistics

Clinical findings were tallied from the EDS1261GW1-23 database, gene and DNA variants from the data in S2-S5 Tables. Tallies used the search, find, and sort functions of Excel, statistical calculations of averages and standard deviations performed using its standard formulae. Significant differences at the p <0.05 level were determined using online resources [65] that compared means by two-tailed t and proportions by N-1 chi-squared tests.

## Results

### EDS patients and clinical findings

Clinical and molecular analyses of the 1899 patients diagnosed with EDS over a 10-year period from 2011 to 2020 are summarized in Table 1 as expanded from a preliminary report [11]. Initial referrals were prompted by complaints of joint pain in 50% and findings of autonomic imbalance in 42%, the remainder by neuromuscular or skeletal complaints with less than 1% referred because of the traditionally emphasized joint hypermobility and skin changes [4–6]—see the Supporting Information Appendix and S1 Table for details on the presenting complaints and patient findings. Thus the conjunction of joint-skeletal and autonomic symptoms emphasized in this article was not an artifact of referral, patients with either complaint having substantial findings of each.

As author interest and the focus of a private setting attracted more EDS referrals, under-appreciated symptoms like brain fog, chronic fatigue, or bowel irregularity [9–11] were recognized and incorporated into a systematic assessment for 80 history and 40 physical findings that included 36 consensus criteria for EDS [66]. Emphasis on overall EDS finding pattern and its relation to underlying tissue laxity-autonomic mechanisms guides the unique approach to DNA variant interpretation in this and a preliminary article [2].

Average numbers of Hx (history) or physical (PE) findings for the 1261 EDS patients meeting criteria [66] and having systematic eval (evaluation) for S1 Table findings were significantly higher (p <0.05) than those for **a**EDS males; bEDS females under age 10.5 years, and cNo EDS patients; DNA all, patients with DNA testing, DNA+, those with a variant reported; X^-^±SD, mean plus standard deviation.

Findings in S1 Table, appreciated in the first 638 patients who met EDS criteria [66], are grouped in 12 history and 7 physical categories. This allows comparison of total numbers of findings, numbers in a category, or individual finding percentages among EDS groups. Despite large standard deviations, the 34 ± 10 of 80 and 18 ± 4.7 of 40 average findings of 1261 patients diagnosed with EDS (first column, Table 1) were significantly higher than those of 64 who were not (No EDS--7.2 ± 1.3 of 80, 7.6 ± 1.3 of 40, last column). The latter group is an imperfect control since their average age (18 **±**13 years) and percentage of females having systematic evaluations (52%) were lower (data not shown) than those diagnosed with EDS. See the Appendix discussion to appreciate that 200 (16%) of the 1261 EDS patients were related, 11 (18%) of the 62 not meeting EDS criteria having relatives who did.

Greater female severity is shown by higher numbers of history (36 ± 9.8 versus 26 ± 8.0) and physical findings (19 ± 4.5 versus 17 ± 4.8) in 1064 females versus 197 males in Table 1, columns 2-3. The 85% preponderance of females likely reflects intrinsic flexibility (Beighton scores averaging 6.9 maneuvers of 9 versus 5.6 for men, S1 Table) and not selective referral as discussed in the Appendix. Although males with their earlier and more strenuous sports participation tend to present for evaluation at younger ages (21 versus 30 years for females in Table 1), their greater muscle development and lesser flexibility underly their lesser frequencies of most findings (tall stature and pectus among the 15 exceptions in S1 Table).

Moving beyond the joint hypermobility and skin elasticity emphasized by dermatologists Ehlers and Danlos [11] are S1 Table assessments of neuromuscular symptoms like migraines or poor balance that affect a respective 60% or 61% of EDS females, 96% of all patients having at least one of 12 neuromuscular findings by history. Equally frequent are the brain fog (83%) or chronic fatigue (87%) of postural orthostatic tachycardia syndrome (POTS) in EDS females, the bowel irregularity (82%) or bloating-reflux (79%) of irritable bowel syndrome (IBS), and the rashes (42%) or asthma-dyspnea (49%) of mast cell activation syndrome (MACS) in S1 Table. All but 3 of the 1261 patients had 1 of these dysautonomia findings and the average among females and males of all ages was 12 of the 20 findings.

The neuromuscular and dysautonomia findings are inextricably linked to those of joint-skin-vessel laxity as shown in the Appendix, 483 females over 10.5 years who presented with joint pain having similar numbers of joint 5.8 to 5.1 of 10, skin (2.5 each of 10), or dysautonomia findings (12.2 to 13.1 of 20) as the 358 who presented with postural orthostatic tachycardia syndrome (POTS). Underappreciation of these neuromuscular and autonomic problems and their exclusion from consensus findings [66] is a major reason for the diagnostic delays for males (7 years, 21-14) and females (13 years, 30-17) in Table 1, columns 2-3 rows 4-5); their recognition is also crucial for timely diagnosis of younger patients who have yet to develop wear-and-tear joint injuries: Note the fewer history-physical findings in females under age 10.5 years than those of older females (Table 1 columns 4-5, rows 6-7), the reason why only the latter were used for later comparison of EDS patients with different gene changes.

Hypermobility measured by Beighton score [67] plus skin elasticity [68] are external indicators of EDS-dysautonomia pathogenesis, the lesser fleshy constraint of more distensible vessels decreasing blood return to the brain with adrenergic reaction. More joint motion leads to wear-and-tear injuries (sprains-- 56% of women, ligament tears-36%, fractures--49%, stretch marks--59%, scars—43% and skeletal bends [69] or deformities (scoliosis—25% or flat feet—46%) necessarily joined by consequences of flexible tissue in other organs (see S1 Table for finding frequencies). Flexible glia, dura and vertebrae allow descent of the lower brain to produce Chiari deformation in 14% of women [70] plus back pain from spinal disc herniation or degeneration in 42% [13].

Equally flexible pelvic ligaments (exaggerated in women for parturition) lead to pelvic congestion [71–73] and urogenital problems (menorrhagia—67%, endometriosis—33%) that when counted give women an average 1.5-point excess in total history scores (S2 Table). Inflicting circulatory imbalance is the increased distensibility of vessels that leads to lower body blood pooling (57% of EDS women have foot discoloration upon standing), the reactive adrenergic stimulation producing stress-related psychiatric symptoms [74] that combine with those from pain [75–76] and inflammation [77–78]. Some of these changes including rare joint contractures [79] were undoubtedly related to aging as 74 EDS females and 6 males were over 50 years old.

Comprehensive assessment of dysautonomia findings in EDS [9-10, 30-33] is crucial for diagnosis and for correlating its cardiovascular, immune, and inflammatory changes with the gene changes in S2 and S3 Tables. Holistic ascertainment renders EDS types as subtle variations on a theme or spectrum, Appendix discussion indicating that 332 (26%) of 1261 EDS patients met criteria for classical [M130000--6, 66] and 892 (71%) for hypermobile [M130020--4, 66] EDS based on the S1 Table findings. Besides their similar average numbers of total history-physical findings (37-18 for classical, 34-19 for hypermobile EDS patients), all other category numbers or finding proportions were similar except for the presence of unusual scars in 86-92% of classical patients on history-physical versus in 25-21% of hypermobile patients (see Appendix). No EDS patients were encountered with facial changes, bowel/ vessel ruptures, and lethal pregnancy complications of vascular EDS [7] or Marfan syndrome [80], patients with the distinctive habitus, aortic dilation, and eye findings of the latter syndrome excluded from this study.

Perhaps casting doubt on the Fig. 1 qualification of DNA results as relevant to EDS findings are the similar numbers of history-physical findings (35-18) among EDS patients having DNA variants (DNA+ in Table 1) as those without (DNA-, 35-19). The most likely explanation for this is incomplete discovery of EDS-contributing genes, prior exome studies finding 9 variant genes in 177 patients [1], 4 in 59 patients [3] and even the present 330 variant genes in 568 patients (317 relevant to EDS) not including mutations outside of exons or exon-intron borders. Support for EDS-DNA correlation by the Fig 1 protocol will follow its discussion and include differences from disability patient results (S4 Table) and the recurring gene variants of S2, S3 Tables.

### A novel clinical protocol for DNA variant qualification

The novel qualification protocol in Fig 1 was developed to add biochemical and clinical considerations to qualification of the average 12,000 DNA sequence changes found in the typical exome [81]. Sophisticated analysis by pioneering laboratories [61–62] developed filters for those DNA variations likely to correlate with patient findings yet crossing thresholds from individual characteristics to the finding patterns of diseases like EDS has proved challenging.

The stepwise protocol in Fig 1, modified from prior publications [2, 82], begins with consensus qualifications [83–84] of pathogenic, likely pathogenic, or variants of uncertain significance based on conformational grading of product disruption [D-85], evolutionary conservation of the altered gene region [E-86], functional *in silico* analysis [F-87] and the dynamic G that increases or decreases as a DNA variant is detected in similarly affected [88–89] or normal individuals [89–91].

Clinical steps are added (columns 3-5 of Fig 1) to consider abundant disease-related symptoms (H), inheritance (I) from relatives with these symptoms and whether the additional (adjunct-J) variants act by synergistic (S) or other (O) mechanisms based on prior disease associations [92, see S2 and S5 Tables]. Each DNA variant is assigned evidenced (VEDU), strong (VSDU), moderate (VMDU) to uncertain (VUDU) or no (VNoDU) diagnostic utility (column 4, Fig 1, each patient DNA result of one or more variants assigned 1-4+ MDna medical diagnostic utility (last column).

The additional clinical correlation emphasizes the entire profile of disease (i. e., all skin-skeletal, neuromuscular, and dysautonomia findings of an EDS patient) rather than a single one like kyphoscoliosis [93], an approach essential for relating syndromic pattern to mechanism. Thus mitochondrial DNA polymerase gamma (*POLG*) variants [27] would be related to the developmental disability of patient 4 (Fig 1, last column) based on that disease association (M302700+) but to dysautonomia symptoms of the 17 EDS patients in S2 Table based on the encephalopathic-gastrointestinal dysmotility symptoms of its other associated disease (M613662+). Changing “molecular” diagnosis [92] to diagnostic utility, minimizing use of “uncertain significance,” and adding qualifiers with connotations like VUDU, VnoDU, V*DUO (dual) in Fig. 1 could lessen physician skepticism [94] and make clear that the most established molecular change may not match clinical symptoms as shown by sickle cell anemia [95]—see the Appendix for additional discussion of the example patients in Fig. 1 and the Discussion for further comments on the relation of DNA variation (as relevant to EDS) to clinical expression (as EDS types or other connective tissue dysplasias).

### Different implications of DNA variants in EDS and disability patients

Support for clinical-DNA correlation in EDS patients will now be provided by comparing their DNA results with those of developmental disability (DD) patients in Fig 2 (see S4 Table for all 167 DNA variants in the 82 disability patients, 13 of them copy number variants). The 967 EDS patients with DNA testing shown in Table 1 are further broken down into 906 having whole exome sequencing and 61 having gene panel or allele testing in Table 2, commercial laboratories reporting potentially significant DNA variants in 568 patients, 32 of them from allele-panel testing. All but 6 results are from the GeneDx© Company as indicated in the S3 list of DNA variants in EDS patients.

**Table 2.** DNA testing results in patients with EDS and developmental disability a. Gene panels were performed on 31 EDS patients (18 with variants, all systematically evaluated), allele testing on 30 EDS relatives (19 with variants, 14 systematically evaluated); **b**9 had allele testing, 4 with potentially significant variants but not meeting EDS criteria; **c**459 had microarray analysis, 102 (22%) having copy number variants including 11 of the 76 with positive WES, 6 of the 82 had positive panel testing (S5 Table); **d**2 patients had incidental variants [60]; **e**All 82 patient results were qualified with <u>diagnostic utility for DD (developmental</u> <u>disability)</u> since that was the indication for testing;*significantly different (p <0.05) from EDS (see Methods); LkPath, likely pathogenic; Path, pathogenic; WES, whole exome sequencing.

| Patient groups | EDS All | EDS Females | EDS Males | Not EDS | DD |
| --- | --- | --- | --- | --- | --- |
| Patients diagnosed with EDS or developmental disability DD = P | 1899 | 1553 | 346 | 80 | 735 |
| Patients having--systematic evaluations (% of P) | 1261(66) | 1064(69) | 197(57)* | 64(80) | --- |
| --DNA testing ((% of P) | 967(51)<sup>a</sup> | 816(53) | 151(44)* | 23(29)*<sup>b</sup> | 461(63)<sup>c</sup> |
| --WES testing (% of P) | 906(48) | 777(50) | 129(38)* | 14(18)* | 112(15)* |
| --a potentially significant DNA variant (% of those having WES) | 536(59) | 459(59) | 77(60) | 0* | 76(68)* |
| --a potentially significant variant by WES, panel, or allele testing = V | 568<sup>d</sup> | 480 | 88 | 4 | 82<sup>e</sup> |
| --at least one LkPath/Path DNA variant for EDS *or* DD by lab (% of V) | 20(3.5) | 16(3.3) | 4(4.5) | 0 | 48(59) |
| --at least one LkPath/Path DNA variant for other diagnosis by lab (% of V) | 181(32) | 154(32) | 27(31) | 0 | 12(15) |
| --variant(s) with 4+ diagnostic utility for EDS *or* DD by author (% of V) | 414(73) | 358(74) | 58(66) | 0 | 53(65) |
| --variant(s) with 3+ diagnostic utility for EDS *or* DD by author (% of V) | 122(22) | 93(19) | 27(31) | 3(75) | 27(33) |
| --variant(s) with 2+ diagnostic utility for EDS *or* DD by author (% of V) | 29(5.1) | 26(5.4) | 3(3.4) | 1(25) | 2(2.4) |
| --a primary mitochondrial DNA variant (% of V) | 93(16) | 79(16) | 14(16) | 0 | 4(4.8)* |

The Fig 1 protocol qualified the variants or variant combinations in all but 2 EDS patients with incidental findings as having utility for that diagnosis (column 1 of Table 2, one of the latter (patient 567 in Table S3) possibly relevant with 15q13 microdeletion that encompassed the CHRNA (M100690) cholinergic receptor gene (M100690). Relating results to tissue laxity-dysautonomia mechanisms qualified 414 or 73% of patients as having results of 4+ medical diagnostic utility and 122 or 22% as having 3+medical diagnostic utility for EDS (Table 2, column 1). Only 20 (3.5%) of patient results were accorded likely or full pathogenesis for EDS-like disorders in commercial laboratory reports, 181 (32%) judged pathogenic for other diseases and 367 (568 minus 201 or 65%) given unhelpful qualifications as variants of uncertain significance. See the discussion of Fig 1 example Patient 5 in the Appendix for the difference in approach to qualification of the 28 primary and 12 additional profilaggrin (*FLG*-M135940+) gene variants in S2 Table), commercially reported as pathogenic for scaly skin (M146700) but here viewed as contributing to skin laxity and adrenergic-inflammatory excess [8, 19, 25–34].

Quite different were the diagnostic implications of DNA results in 82 patients with developmental disability, all of them related to the latter indication for testing and some to specific disability syndromes [S5 Table]. It must be noted that 49% of disability patients were female (not shown) compared to the 85% of EDS patients in Tables 1-2 and that whole exome sequencing was usually performed in disability patients after microarray analysis was normal [63]. Of the 459 having microarray analysis, 102 (22%) had potentially significant copy number variants including 11 of the 76 with positive whole exome sequencing (last column, Table 2). In contrast, only 9 EDS patients had copy number variants found by simultaneous testing [64] with 3 judged relevant to EDS (Table S3).

The 330 genes variant in EDS patients and their prior disease associations are listed in S2 Table, M numbers from www.omim.org provided as references. The 10 genes with 13 DNA variants not considered relevant to EDS are at the bottom along with 3 genes and 5 variants considered incidental or secondary findings [60]. The 917 DNA variants in 568 EDS patients are listed in S3 Table by patient numbers, single and therefore primary variants having .0 after the patient number, multiple variants followed by .1 for the one judged primary and 0.2, 0.3, etc. for additional variants (modeled by the example variants of Fig 1, last column). When two or more variants occur in the same gene, they are given separate numbers and labelled as homozygous (18 variants, 9 patients), trans (47 variants, 23 patients), cis (23 variants, 12 patients), ?cis-trans (10 patients) or cis + trans (patient 231 with 3 variants) in column D of S3 Table. Of the 911 DNA sequence variants cited in commercial reports, 561 (62%) were listed in ClinVar and 71 of 158 mitochondrial DNA variants (45%) were listed in MitoMap (see legend to S3 Table).

Patients are numbered from low to high according to how much their altered genes are thought to contribute to EDS, those with variants in well-recognized genes like collagen type V [6] having low numbers and those given novel relevance by this study (e. g., collagen type VI [19] or mitochondrial ATP synthase [24] variants) having higher numbers. The disability variant list in S4 Table is similarly numbered and qualified but ordered by date of entry since all were relevant to developmental-intellectual disability.

There were 20 genes (with 24 variants in 21 disability patients) that were also variant in EDS patients (blue colors in S3 and S4 Tables). Nuclear genes include *ATP7A*, *DUOX2, FLNA*, *POLG*, and *TG*, mitochondrial ones *MT-CO2*, *MT-TK*, and *MT-ND5*, their different mutations feasibly contributing to cognitive disability on the one hand or to the autonomic and neurologic issues of EDS on the other. Also in both patient groups were the profilaggrin gene *(FLG*, M135940) variants discussed above, present in 2 (2.4%) of the 82 disability and 35 (6.2%) of the 568 EDS patients (S2 and S4 Tables). Only in the latter group was their prevalence more than the 2.2% in normal individuals [90, 91], supporting their autonomic-inflammatory action in some EDS patients. Additional variants in the connective-tissue related *COL11A*, *PLOD1*, and *FBN2* genes in disability patients may augment the hypermobility often related to low muscle tone as reported in a child with Down syndrome [96].

Also supporting clinical correlation are the DNA variant distributions shown in the upper rows of Table 3, 221 or 39% of EDS patients having 327 additional variants and 43 or 52% of disability patients having 72 of them. The latter total does not count the 12 chromosome or copy number variants that may contribute to patient disabilities, the 17q21.31 microdeletion in patient 33 of S5 Table likely contributing more than its companion *G3BP1* (M608431) gene sequence alteration to disability yet rated primary to aid EDS-disability variant comparisons.

**Table 3.** Qualification and parental origin of DNA variants found in EDS or disability patients. ^a^566 patients had EDS-relevant primary DNA variants including 345 (61%) with one and 221 (39%) with one primary and 327 additional variants, 221 (68%) of those having one additional, 77 (24%) two, 22 (6.7%) three, 6 (1.8%) four, and 1 (0.31%) five. ^b^82 had primary DNA sequence variants, 39 (48%) with one and 43 (52%) with one primary and 72 additional variants, 43 (60%) of those having one additional, 20 (28%) two, 8 (11%) three, and 1 (1.4%) four; ^c^unknown--no parental samples, undetermined--inability to distinguish *de novo* or maternal origin; ^d^percentages in these rows refer to proportions of all DNA variants;* significant (p < 0.05) difference between EDS and developmental disability (DD) patients (see Methods).

| Categories of DNA variants | EDS<br>All | EDS<br>Primary | EDS<br><u>Additional</u> | DD<br>All | DD<br>Primary | DD<br><u>Additional</u> |
| --- | --- | --- | --- | --- | --- | --- |
| **Total DNA variants T** | **893<sup>a</sup>** | **566<sup>a</sup>** | **<u>327<sup>a</sup></u>** | **154<sup>b</sup>** | **82<sup>b</sup>** | **<u>72<sup>b</sup></u>** |
| VEDU or <u>VEDUS</u> (%T) | 384(44)* | 359(63) | <u>29(8.9)</u> | 61(35)* | 53(60) | <u>3-8(11)<sup>c</sup></u> |
| VSDU or <u>VSDUS</u> (%T) | 321(36)* | 170(30) | <u>151(46)</u> | 69(60)* | 27(38) | <u>32-42(58)</u> |
| VMDU or <u>VMDUS</u> (%T) | 169(19)* | 35(6.2) | <u>134(41)</u> | 24(4.7)* | 2(2.4) | <u>9-22(30)</u> |
| VUDU or <u>VUDUS</u> (%T) | 15(1.7) | 2(0.35) | <u>13(4.0)</u> | 0 | 0 | <u>0</u> |
| **Nuclear variants N(%T)** | **735(82)\*** | **473(84)\*** | **<u>262(80)</u>** | **144(93)\*** | **78(95)\*** | **<u>66(88)</u>** |
| maternal origin (%N) | 245(33)* | 163(35)* | <u>82(31)</u> | 30(21)* | 13(17)* | <u>17(26)</u> |
| paternal origin (%N) | 193(26) | 122(26) | <u>71(27)*</u> | 42(29) | 16(21) | <u>26(39)*</u> |
| *De novo* (%N) | 31(4.2)* | 18(3.8)* | <u>13(5.0)*</u> | 48(33)* | 35(45)* | <u>13(20)*</u> |
| Unknown<sup>d</sup> (%N) | 266(36)* | 170(36)* | <u>96(37)*</u> | 24(17)* | 14(18)* | <u>10(15)*</u> |
| **Mitochondrial variants M(%T)** | **158(18)\*** | **93(16)\*** | **<u>65(20)</u>** | **10(6.5)\*** | **4(4.8)\*** | **<u>6(12)</u>** |
| maternal origin (%M) | 102(65) | 58(62) | <u>44(68)</u> | 6(60) | 2(50) | <u>4(67)</u> |
| paternal origin (%M) | 0 | 0 | <u>0</u> | 0 | 0 | <u>0</u> |
| *De novo* (%M) | 4(2.5) | 4(4.3) | <u>0</u> | 0 | 0 | <u>0</u> |
| Undetermined<sup>c</sup> (%M) | 52(33) | 31(33)* | <u>21(32)</u> | 4(40) | 2(50)* | <u>2(33)</u> |

The importance of additional variants that are often ignored in published work is shown by the 96% in EDS and 44% in disability patients that are qualified with moderate to evidenced synergistic contribution (V-E/S/M-DUS in Fig. 1, columns 3 and 6 of Table 3). Note that 28 or 39% of the 72 additional variants in disability patients were associated with other diseases (column 6, Table 3) compared to the 13 or 2.0% of 911 in EDS patients listed at the bottom of S2 Table that are not included in the Table 3 totals. The latter list may expand when large variant databases become available although the 143 genes represented by multiple variants will likely achieve evidenced diagnostic utility for EDS.

The lower rows of Table 3 show dramatic differences in origin of variants in EDS versus disability patients, primary nuclear variants having maternal origin in a statistically significant 35% versus 17% while *de novo* variants have the reverse difference of 3.8 versus 45% that is also significant (see Methods). Mitochondrial variants are much more prevalent in EDS patients (bottom of Table 2, reiterated in Table 3, mapped in Fig. 2B), surprising in view of their associations with severe disability diseases such as Leigh syndrome (M256000). The 93 primary and 65 additional mitochondrial DNA variants in EDS, like others associated with neurologic disorders, emphasize the importance of muscle in protecting joints and constraining tissue-vessels to promote blood return to the upper body and brain.

### An EDS gene network

The 65 genes with 4 or more variants in EDS patients have a broad distribution in the nuclear genome (Fig 2A—bold*, red print), matched by 30 genes in the mitochondrial genome (Fig 2B) where the DNA/protein changes are specified. With less certain relevance but equally wide distribution are the 252 genes with fewer than 4 variants (bold*, black print in Fig 2A), 110 of them with no primary and only additional variants (italic+, black print in Fig 2A, filled black squares in Fig 2B).

Genes are classified by their impact on tissue elements (e. g., joint. Jt) or processes (e. g., Ans, general autonomic regulation) according to their previous associations with disease as shown in the lower box of Fig 2A and the legend of S2 Table; these classifications are listed beside the genes with primary variants in Fig 2A. Variants in nuclear genes that encode products routed to the mitochondrion are in green print in Fig 2A and listed for the mitochondrial gamma polymerase (*POLG*) gene in Fig 2B, its role in mitochondrial depletion with the associated neuromuscular (M607459+) and dysautonomia (M612662+) conditions discussed above important for understanding how mitochondrial dysfunction might contribute to EDS.

Another classification in S2 Table important for later EDS-COVID19 comparisons pertains to the nature of the RNA or protein product encoded by the gene, terms like Ez for enzyme, Mc for membrane channel, or Tf for transcription factor explained in that Table legend. The transcription factor group includes 26 or 8.2% of the 317 genes relevant to EDS (S2 Table) and suggests that many EDS-relevant mutations in regulatory regions outside of exon or exon-intron borders remain to be discovered. The diverse element-process impacts and products of EDS genes are paralleled by their diffuse genomic locations, clustering evident only for *COL5A2/COL3* at 2q32.2, *SCN5/10/11A* at 3p24.1, *COL6A1/A2* at 21q22.3, and *SCN2B/4B* at 11q23.3.

### Similar clinical profiles in EDS patients with different gene changes

EDS patients with multiple variants in the same gene are shown in Fig. 3, extracted from the larger list in Table S2 that provides complete gene names, M number references, and associated diseases. Key groups in the upper rows include 51 patients with primary collagen type V variants, 35 in the *COL5A1* gene, 32 in female patients over 10.5 years with sufficient EDS-dysautonomia findings for comparison (see Table 1). Because of their long and accepted association with EDS and their numbers, patients with *COL5A1* gene variants were chosen as a reference for the other patient groups, few significant differences (squares, circles) noted in categories ranging from total history to total dysautonomia findings in Fig 3.

While collagen type V gene variants were previously associated with classical EDS [6] and collagen type III variants (13 primary, 12 qualifying for comparison) with vascular EDS [7], these patients had very similar numbers of tissue laxity, neuromuscular, or dysautonomia findings (colored columns of Fig. 3). The same goes for patients with collagen type I variants, usually associated with osteogenesis imperfecta but recently with an EDS phenotype [98]. Similarity of clinical profiles extends to patients with variants in the fibrillin-1 (*FBN1-*-13 qualifying patients) and transforming growth factor/receptor genes (*TGFB/TGFBR*–11 qualifying patients) that were previously associated with the connective tissue dysplasias Marfan (M154700) or Loeys-Dietz (M609192+) syndromes. The latter patients’ compatibility with EDS is reaffirmed by the exclusion of patients with the obvious clinical diagnoses of Marfan or Loeys-Dietz syndromes from this study (see Methods). Patients with *COL3* and *FBN1* gene changes, reflecting association of other mutations in these genes with severe disease, did have significantly higher numbers of findings in the total history-physical and certain other categories depicted in Fig. 3.

One can further relate the collagen type I, III, and V genes to impact on particular tissue elements via their associated diseases and tissue distribution, I with major relation to bone [99], III to vessel and other hollow organs [7,100], and V to joints [97] as indicated in the lower box of Fig. 2A and the legend to S2 Table. Similar relations of collagens VII [101] and XVII genes to skin (5 patients qualifying for comparison), collagen IX genes to bone (8 patients), and the von Willebrand factor gene [17, 20] to vessel lining/clotting (7 patients) complete the tissue laxity groups in the upper rows of Fig 3. Note that other tissue laxity groups have similar numbers of tissue laxity, neuromuscular, and dysautonomia findings as the patients with *COL5A1* gene changes although the *COL7/17* patients have a significantly higher average Beighton score and *VWF* patients significantly more physical findings.

The congruence of finding profiles continues with the middle and lower patient groups of Fig 3, COL6 (9 qualifying) or COL12 patients (13) with genes impacting muscle, the several groups with mitochondrial gene variants (19, 12, 9, and 15 patients), and the sodium channel SCN9A patients (7 qualifying) having few significant differences. A different way of grouping patients is exemplified by those with variants in genes like *AARS1* that have been associated with forms of Charcot-Marie-Tooth disease and therefore impact peripheral nerves (Np). These 22 patients with variants in 17 genes (Table S2), 13 with primary variants and 10 qualifying for comparison, are the only group with genes of neuromuscular impact that have significantly more neuromuscular findings in Fig 3.

The lower rows of Fig 3 also group genes by impact on disease process, the 8 qualifying patients with porphyria-associated genes like *HMBS*, the 17 with mitochondrial *ATP6* gene variants, the 14 and 23 patients with variants in the previously discussed *POLG* [27] and *FLG* [102] genes again having similar finding profiles. However, the latter two groups with exaggerated dysautonomia and inflammatory impact--judged from their associated sensory-bowel immotility (M613662+) and atopic predisposition (M605803)--had significant differences in 4-5 categories.

Not shown are considerable data comparing frequencies of individual findings, a few trends emerging like more tall stature/angular build in patients with the Marfan-related *FBN1* gene variants. This additional data also finds few significant differences among finding proportions in gene groups, supporting network action in normality and disease but needing more DNA findings for firm conclusions.

### Novel and interesting genes showing variation in EDS patients

Continuing the theme of genes being part of a network contributing to general EDS-dysautonomia findings are the many genes of interest in Table 4, categorized by impact on tissue element or process as described in the lower box of Fig 2A. Key points illustrated by these genes are presented here; more details and many other EDS-related genes in each category are discussed in the Appendix.

**Table 4.** EDS-related genes of interest. Genes asimilar or didentical to those influencing COVID19 severity (S5 Table); balso found in disability patients (S4 Table); cassociated disorders not having 3 or more findings of EDS; gene names and additional list of associated disease ymptoms in S2 Table; symbols for tissue elements or processes impacted in Fig. 2a box; AD, AR—autosomal dominant, ecessive; CMT, Charcot-Marie-Tooth; IBS/IBD, irritable/inflammatory bowel disease; ID, intellectual disability; MVP, mitral valve prolapse; XL, X-linked.

| Gene | El-P | PV | AdV | Associated diseases and selected findings (EDS-related are underlined and italicized) |
| --- | --- | --- | --- | --- |
| **Tissue dysplasia** |  |  |  |  |
| *ABCC6* | Jt | 3 | 0 | Pseudoxanthoma elasticum, form fruste M177850+ AD—brain hemorrhage, vascular calcification, atherosclerosis |
| *ADAMTS2<sup>a</sup>* | Jt | 2 | 0 | “EDS dermatospraxis type” M225410+ AR—short stature, *skin elasticity*, *myopia*, *hypodontia*, *pneumothorax* |
| *FKBP14* | Jt | 2 | 0 | “EDS kyphoscoliotic type-2” M614557 AR—blue sclera, *skin elasticity*, *scoliosis*, *fractures*, *myopathy*, *bladder diverticula*, *hernias* |
| *PLOD1<sup>b</sup>* | Jt | 5 | 1 | “EDS kyphoscoliotic type-1” M225400 AR—*Marfanoid habitus*, *myopia*, *retinal detachment*, *scoliosis*, *weakness* |
| *TNXB* | Jt | 7 | 3 | “EDS classic-like-1” M606408+ AR—*joint pain*, *soft-lax skin*, *MVP*, *aortic valve anomaly*, *bladder issues*, *weakness* |
| *FLNA<sup>b</sup>* | Jt | 3 | 4 | Periventricular nodular heterotopia 1 M300049+ XLD—*aortic dilation*, *respiratory difficulties*, clotting disorder, *bowel dysmotility* |
| *FLNB* | Jt | 1 | 1 | Larsen syndrome M150250+ AD—short stature, *joint dislocations*, *scoliosis*, congenital hip dislocation, hearing loss |
| *FLNC* | Vs | 2 | 0 | Cardiomyopathy, hypertrophic-26 M617047+ AD—*MVP*, *arrhythmia*; Myopathy distal 4 M614065 AD—*weakness* |
| *ZNF469* | Jt | 2 | 2 | Brittle cornea syndrome-1 M229200 AR—corneal ruptures, Marfanoid habitus, *blue sclerae*, *unusual scars*, *lax skin* |
| *F10* | Clot | 2 | 1 | Factor X deficiency M227600 AR—nose and *gum bleeding*, *easy bruising*, hemarthroses, *menorrhagia* |
| *KCNA5* | Vs | 1 | 0 | Familial atrial fibrillation-7 M612240 AD—*arrhythmia*, atrial fibrillation, *palpitations*, *fatigue*, *exercise intolerance* |
| *LOX* | Vs | 3 | 0 | Aortic aneurysm, familial thoracic-10 M617168 AD—*skin striae*, *tall*, *pectus*, *hernia*, *dural ectasia*, *MVP* |
| **Developmental** |  |  |  |  |
| *SKI* | Vs | 1 | 0 | Shprintzen-Goldberg syndrome M182212 AD—lax skin, MVP, aortic dilation, asthma, bowel dysmotility, Chiari, ID |
| *LIFR<sup>d</sup>* | Bn | 0 | 1 | Stuve-Wiedemann/Schwartz-Jampel type 2 syndrome M601559+ AR—*feeding*, *lucent skin*, *scoliosis*, *dysautonomia* |
| *NOTCH1<sup>c</sup>* | Vs | 2 | 0 | Adams-Oliver syndrome-5 AD M616028—brain anomalies, blood clots, bicuspid, stenotic aortic valve |
| *MYBPC3<sup>c</sup>* | Mu | 3 | 5 | Cardiomyopathy, dilated M615396+ AD—cardiomyopathy, arrhythmia-ventricular flutter |
| **Neuromuscular** |  |  |  |  |
| *MED12* | Nc | 2 | 0 | Lujan-Fryns syndrome M309520 XLR—*tall*, *high palate*, *angular*, septal defects, aortic *aneurysm*, *pectus*, ID-autism |
| *ATP7A<sup>b</sup>* | Nm | 1 | 1 | Occipital horn syndrome M304150+ XLR—exostoses, *elastic skin*, *pectus*, carotid tortuosity, *bladder diverticula*, ID |
| *LMNA* | Nm | 1 | 0 | CMT type 2B1 M605588+ AR—*CMT symptoms*, *kyphoscoliosis*; Pprogeria M176670 AD—*lucent skin*, *aging*, etc. |
| *MT-CO2<sup>b</sup>* | Nm | 3 | 1 | In patients (M516040) with *hypotonia*, *ataxia/poor balance*, optic atrophy, *cardiomyopathy*, *fatigue*, *muscle weakness* |
| *CLCN1<sup>c</sup>* | Mu | 1 | 2 | Myotonia congenita, dominant M160800+ AD—muscle stiffness, *muscle aches*, eyelid/tongue myotonia |
| *LPIN1* | Mu | 2 | 2 | Myoglobinuria, acute recurrent M268200 AR—*weakness*, *muscle aches*, rhabdomyolysis, *stress/fever-induced* |
| *CACNA1H* | Ans | 2 | 1 | Hyperaldosteronism, familial, type IV M617027+ AD—hypertension, *muscle weakness*, epilepsy susceptibility |
| *TMPRSS6<sup>ac</sup>* | Ans | 0 | 1 | Iron-refractory iron deficiency anemia M206200 AR—*bowel issues* with iron malabsorption, iron deficiency anemia |
| **Dysautonomia-inflammation** |  |  |  |  |
| *RYR2* | Mu | 3 | 1 | Ventricular tachycardia, adrenergic polymorphic-1 M604772+ AD—*arrhythmia*—exercise/*stress*-induced; *syncope* |
| *SCN11A* | Ns | 4 | 0 | Hereditary sensory-autonomic neuropathy VII M615548+ AD—*bowel dysmotility*, *reactive skin*, *weakness*, *fractures* |
| *HFE* | Apor | 3 | 13 | Porphyria susceptibility M176100/176200+ AD—*bowel dysmotility*, *neuropathy*; Hemochromatosis M235200 AR. |
| *NTRK1* | Ans | 2 | 2 | Hereditary sensory-autonomic neuropathy V M608654 AR—fevers, *fractures*, *scoliosis*, *dislocations*, *dysautonomia* |
| *SLC6A2<sup>a</sup>* | Ans | 3 | 1 | Orthostatic intolerance M604715—high norepinephrine—*dizziness*, *syncope*, *tachycardia*, *fatigue*, *brain fog*, *anxiety* |
| *NLRP12* | Aim | 4 | 3 | Familial cold autoinflammatory syndrome 2 M611762 AD—*arthritis*, *headaches*, *muscle aches*, *IBS*, *hives*, fevers |
| *NOD2* | Aim | 4 | 0 | Blau syndrome M186580+ AD—uveitis, glaucoma, cataracts, *arthritis*, *rashes*, *scars* *contractures*, IBD, fevers |

#### Genes associated with connective tissue dysplasias (Jt)

- Among tissue dysplasia or joint-impacting genes (Jt) are heterozygous *ABCC6* variants in Table 4, qualified as relevant to EDS despite their prior association with autosomal recessive pseudoxanthoma elasticum [103]. Many heterozygous variants are qualified as contributing to EDS in the way that several haplo-deficient enzymes in biochemical pathways can produce metabolic disease through synergistic heterozygosity [104]. Other heterozygous variants associated with recessive EDS types include those in genes *ADAMTS2*, *FKBP14, PLOD*, and *TNXB* of Table 4, all occurring in patients with typical EDS-dysautonomia profiles.
- Variants in filamin A (*FLNA*) at Xq28 associated with periventricular heterotopia [105] or in *FLNB* at 3p14.3 and *FLNC* gene at 7q32.1 associated with different diseases (Table 4) indicate how homologous genes affecting connective tissue could acquire new components and diffuse into different genomic regions. Mutations in homologous rather than divergent domains of these genes could produce similar EDS rather than divergent phenotypes.
- The 26 genes encoding transcription factors like *ZNF469* suggest that whole genome sequencing will find many additional variants in extra-genic regulatory regions, explaining why 41% of EDS patients had no significant variants identified by whole exome sequencing (Table 2).

#### Genes associated with cardiovascular (Vs) and clotting (Clot) functions

- Variants in the *VWF* gene [17,20 Table 3] and that in the *GP1BA* gene also associated with a form of von Willebrand disease (M177820+) show the importance of blood vessel connective tissue for platelet adhesion and clotting. Other EDS-related genes associated with cardiovascular elements include those associated with bleeding (*ABCC6* and *F10* genes in Table 4), vessel dilation (*FLNA*, *LOX*, *SKI*, *MED12*, *ATP7A)* arrhythmias (*FLNC, KCNA5*, *RYR2)* and cardiomyopathy (*FLNC, MT-CO2, MYBPC3, NOTCH1)* in Table 4, the latter genes given the VsCM designation in S2 Table.
- Heart-related genes affecting development include *SKI, NOTCH1,* and *LMNA* in Table 4, the latter associated with multiple diseases including Hutchison-Gilford progeria with rapid aging (M176670). The *TGFB* and *TGFBR* genes of Fig. 3 have been associated with heart defects as well as immunity [34, 106] and wound healing [107] that can be deficient in EDS. Genes contributing to congenital malformations show the need for experienced clinical qualification of their DNA variants since EDS findings like deformations, dilations, or slippages are almost always acquired rather than congenital.
- EDS-associated genes in Table 4 that influence heart-related lipid metabolism include *LPIN1* (Table 4) and COL3 [Fig 3, 16] involved in adipogenesis and muscle disease. Others like *LDLR* (M606945) influencing cholesterol transport are discussed in the Appendix, their associations with muscle perhaps explaining those side effects of statin medications [108].

#### EDS-contributing genes associated with neuromuscular disorders

- Genes altering brain function (Nc) include *FLNA, MED12,* and *ATP7A* [109] in Table 4, respectively associated with the mentioned periventricular heterotopia, Lujan-Fryns, or occipital horn syndromes that have many symptoms of connective tissue dysplasia. A theme worth exploring is action of these genes on the glial connective tissue surrounding CNS neurons as by the transforming growth factor-beta related genes of Fig 3 [34].
- The need to correlate gene changes with articulo-autonomic dysplasia mechanisms is again shown by the fact that periventricular heterotopia and occipital horn syndrome were once considered types of EDS [105, 109]. In turn and as emphasized before, ascertainment of finding patterns rather than of single abnormalities like brain heterotopia is essential for recognizing underlying pathogenic mechanisms.
- At the same time, the difficulty of classifying genes by impact on one tissue element or process as attempted in S2 Table and Fig. 4 below is illustrated by the lamin AC gene in Table 4. It is associated with 10 diseases including those causing heart-brain-genital anomalies (M610140, M212112), cardiomyopathy (M115200), the Hutchison-Gilford progeria syndrome with rapid aging (M176670), and a form of Charcot-Marie-Tooth disease (M605588).
- Grouping of genes by impact on peripheral nerve (Np) does seem appropriate since the consistent phenotype of Charcot-Marie-Tooth disease with its classic steppage and foot-drop gait is associated with 7 genes and their 22 variants including *LMNA* and *AARS1* in Fig. 3.
- Impacting neuromuscular function (Nm) are mineral transporters of sodium (*SCN9A,* others impacting neurosensory (Ns) functions in Fig. 3) and of calcium, potassium, or copper (*CACNA1H*, *KCNA5, ATP7A* of Table 4), the latter previously associated with Menkes (M309400) or Wilson (M277900) diseases [109]. Copper is also important for *LOX* lysyl hydroxylase and the *MT-CO2* component of complex IV [110] in Table 4. Fibulin-4 is required for activation of LOX in the mouse [111], relating two fibulin-like genes (*EFEMP2, FBLN5* in S2 Table) to lax skin diseases (M614437, M614434) with elastin fiber deficiency. Fibulin 5 is also associated with a form of Charcot-Marie-Tooth disease (M619764), these genes linking collagen cross-linking, heart [112], oncologic [113], and neurologic diseases [114] to cupric influence on connective tissue.
- Iron seems involved with EDS via the *TMPRSS6* gene variants associated with iron malabsorption, the *HFE* gene variants [115] conferring porphyria susceptibility, and the *NUBPL* gene variant associated with complex I deficiency (M618242) in Tables 4 and S2. The chloride channel *CLCN1* and the *SLC26A4* gene that transports chlorine and iodine are related to EDS, the latter associated with thyroid dysfunction (M274600) as are 7 other genes in S2 Table. Hypothyroidism is a cause of intellectual disability while hyperthyroidism is associated with autonomic symptoms [tachycardia, bowel irregularity, fatigue, 116-117], both thyroid conditions occurring with COVID19 infection [59]. Although only 5 patients with thyroid-related genes (Athy) qualified for finding comparison (females above 10.5 years) and were not included in Fig. 3, these patients had typical EDS-dysautonomia profiles
- Unexpected in EDS patients were the 158 mitochondrial DNA variants mapped in Fig. 2B and listed in Tables S2, 3 and 4. Those genes variant in EDS include all but 7 of the 37 in mitochondrial DNA, including components of four respiratory chain complexes (I/*MT-ND* genes, III/*MT-CYB* gene, IV/*MT-CO* genes, V/*MT-ATP* genes), the two ribosomal RNAs, and 16 transfer RNAs in Fig. 2B. The diversity of these mitochondrial genes suggests contribution to EDS by depletion of mitochondrial numbers and energy production, a depletion that would most effect active tissues like brain, nerve, heart, and muscle. Mitochondria also have roles in aging [118] and inflammation, the latter emphasized recently in COVID19 sepsis [119] and possibly impacting the mast-cell activation of EDS patients {32, 33].
- Having impact on muscle (Mu) are the many *COL6/12* gene variations in Table 3, two-copy recessive-acting mutations first related to Ullrich muscular dystrophy (M616470), single copy dominant-acting mutations to Bethlem myopathy (616471) and then to EDS [120]. Many genes like the *CLCN1* and *LPIN* genes of Table 4 and 18 others like the myosin heavy chain *MYH2/7/7B/11* genes with variants in 10 EDS patients in Table 2 were previously associated with muscular dystrophies or myopathies.
- Another 68 genes in S2 Table including the *PLOD1*, *TNXB*, and *FLNC* genes in Table 4 are not classified as having primary impact on muscle yet have prominent symptoms of muscle weakness. The many nuclear and mitochondrial gene changes impacting muscle suggest that appropriate exercise will benefit EDS patients in the same way that it lessens similar symptoms in the old [121].

#### EDS-contributing genes associated with dysautonomia and other processes

- Correlating with the many dysautonomia symptoms of Table S1 are 30 genes in Table S2 with general impact on the autonomic nervous system (Ans) including several like *POLG* [27], *MT-ATP6* [24] in Fig 3 and *NTRK1, SLC6A2* in Table 4. Neurosensory impact was exemplified by 7 sodium channel genes with 24 DNA variants in EDS patients (*SCN9A* in Fig 3, *SCN11A* in Table 4, *SCN5A-10A* in Table S2), their actions evidenced histologically by small fiber neuropathy on skin biopsy [12]. Many associated diseases combine the two processes as hereditary sensory and autonomic neuropathies including those associated with the *NTRK1*, *SCN9A*, *SCN11A* (M615548), and *SPTLC2* (M613640) genes in Table 4.
- Five genes in 18 EDS patients are associated with porphyrias (Apor) and their many autonomic symptoms like the ones encoding the mentioned HFE iron regulator [115] that is variant in 9 patients or the HMBS porphobilinogen deaminase (Fig 3, Table 4) that is a key step in the porphyrin synthesis pathway. The 22 DNA variants in these two genes mandate evaluation of tissue laxity symptoms in porphyria diseases that are noted for abdominal pain crises, bowel dysmotility, tachycardia, neuropathies and, in some cases, skin changes (M618892); they also suggest attention to porphyrin metabolism in some patients with connective tissue dysplasias.
- Primary variants in genes like *SLC6A2* (Table 4) that encode the noradrenaline transporter support the idea that autonomic imbalance can increase tissue laxity [2, 11] through influence on its permeating small fiber neurons [14]. Other links between connective tissue and adrenal/muscle function include variants in the *COLQ* gene [22] that encodes a collagen anchor for a cholinergic receptor and the *SERPINA6* gene, its associated cortisol-binding globulin deficiency (M61148) emphasizing that altered cortisol levels can contribute to the fatigue, muscle weakness, and hypo- or hypertension of EDS (S1 Table, column H).
- Calcium channel genes in addition to those mentioned above and in the Appendix are *RYR1* and *RYR2* (Table 4) that encode products involved in muscle contraction [122]. They additionally increase sensitivity to adrenaline to cause arrhythmias with cardiomyopathy (M604772), conditions that likely relate to the hypotension and tachycardia in EDS patients.
- Serving as a transition to genes associated with long COVID19 are the 28 genes involved in the immunity and inflammatory functions that are impacted by autonomic imbalance (Aim). These include the *NLRP1*/*2/12* genes associated with autoinflammatory diseases (M617388, etc.) that involve arthritis, thyroiditis, skin inflammation, sweating changes, and elevated inflammatory markers. These diseases and those caused by the *NOD1/2* [35] genes in Tables 4 and S2 have overlap with EDS findings like the 14% of females with autoimmune markers (S1 Table) and the chronic variable immune deficiency that is occasionally diagnosed in EDS and may affect COVID19 vaccine response [58]. Included in this group is the *C1R* gene encoding a complement component (S2 Table) that has been associated with an inflammatory (periodontitis) type of EDS [M130080, 123]. Here patient 441 of S3 Table with the *C1R* gene variant had the same EDS-dysautonomia profile as the other patients in Fig 3.
- Genes contributing to EDS thus include many related to immunity and inflammation, from those mentioned above to collagen type I that contains an immunoglobulin receptor binding sequence [15] to transformation growth factor genes [34, 107] and even the mitochondrial genes of Fig. 2B that have roles in immune cells [124, 125]. These genes contributing to EDS will now be compared to those influencing COVID19 severity (Table S5).

### Genes conferring susceptibility to severe COVID19 infection

An obvious difference between the genes contributing to EDS and those influencing severity of COVID19 infection [48–53] is the latter’s modulation of viral infectivity in addition to host responses. SARS-CoV-2 infection depends first on the binding of its spike (S) glycoprotein to the angiotensin-converting enzyme 2 receptor (*ACE 2*, M300335), then on cell entry of the glycoprotein complex by clathrin-mediated endocytosis and cleavage by proteases [44]. The favored route is through nasal epithelial cells with cleavage by transmembrane serine protease 2 (TMPRSS2, M602060) or by lysosomal cathepsin L in other cells. When variations in host genes were associated with severity of the viruses’ associated COVID19 disease, it was not surprising that *ACE2* and *TMPRSS2* were implicated in several studies [126, see S5 Table and the more technical references pertinent to COVID19-relevant genes beneath it].

While the *ACE2* and *TMPRSS2* genes are clearly related to viral processes, many other genes in S5 Table are more difficult to classify, highlighted by their presence in pathways activated during response to infection or by their having selective variation in conjunction with certain symptoms or outcomes [127]. Symptoms of acute infection include fever, cough, fatigue, hoarse voice, loss of appetite, and delirium in over 50% of patients, diarrhea, chest pain/shortness of breath, abdominal pain, and anosmia in under 10% [44]. Outcomes varied from asymptomatic illness to the progressive respiratory failure, renal injury, coagulation changes, and eventual multiorgan dysfunction in 15%, many of the latter older, male, and compromised by obesity, hypertension, diabetes, or heart disease [45].

Since several of the post-acute or long COVID19 symptoms discussed below are prolonged versions of the above, and since their timing and severity meriting “long-haul” diagnosis remain ill-defined, it is not clear which virus-host activities are impacted by many of the genes in S5 Table. Nevertheless, the 104 COVID19-related genes were mapped and classified by element-process or type of encoded product as done for the 317 contributing to EDS in S2 Table, their genomic distribution shown in Fig 2A (blue print), their classification, gene, and symptom comparisons shown later in Figs 4 and 5.

**Fig. 5.**
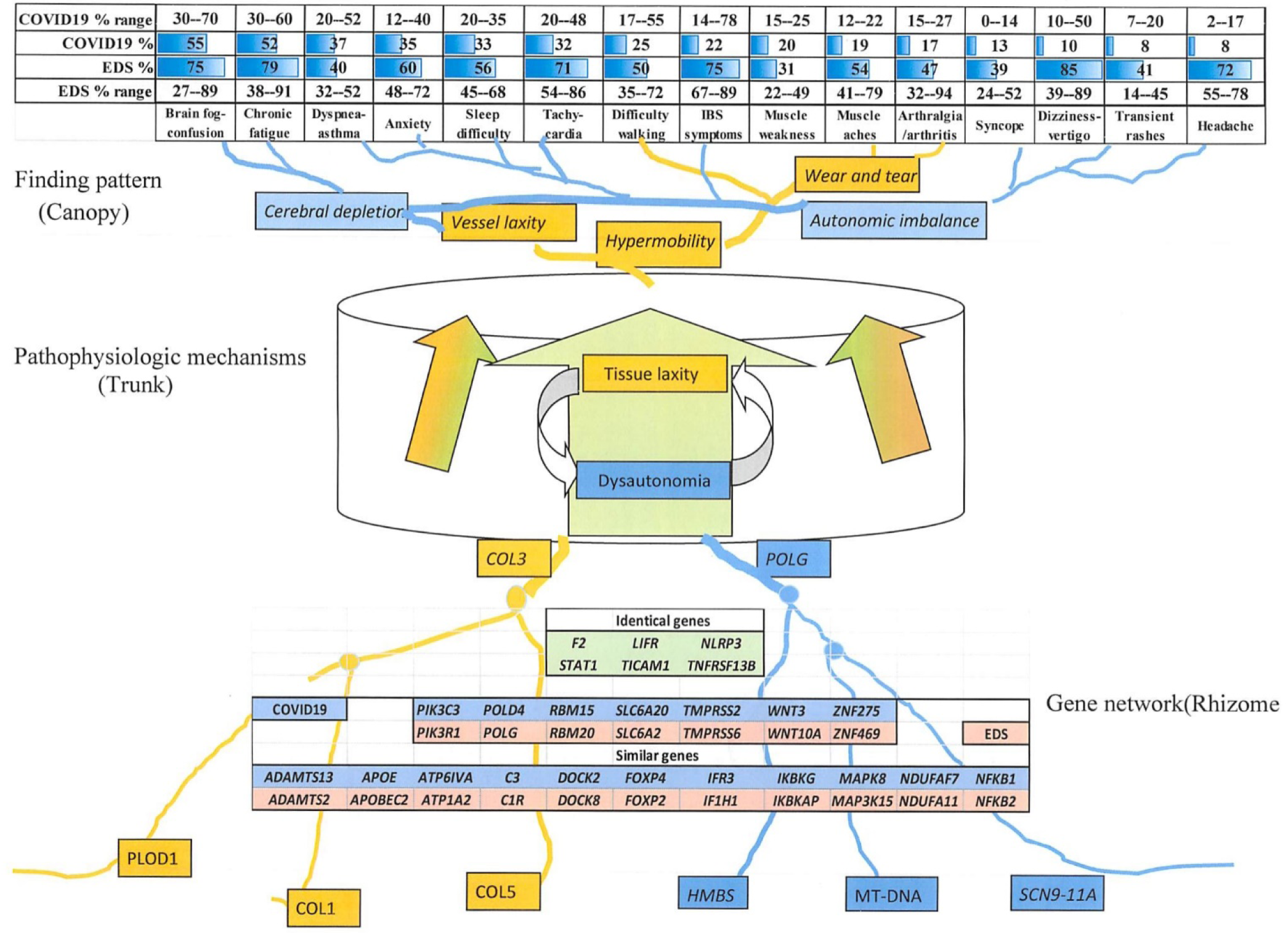
Genes and symptoms related to EDS and COVID19. Genes related to EDS (Table S2) and COVID19 infection (Table S5) are envisioned as overlapping parts of a network (rhizome below) connected through pathogenic mechanisms (trunk sap, phloem) to common symptoms of EDS (Table S1) and long COVID19 (canopy above). EDS symptom ranges are for females over age 10.5 years from the EDS1261database, long COVID percentages and ranges taken from Fig. 2 of Deer et al.[37].

### Comparing COVID19/EDS gene type and distribution

The first thing to note is that the COVID19-relevant genes in Fig 2A are as dispersed as those of EDS except for a *CCR1/5-CXCR6* cluster at 3p22.2. These are chemokine receptors that mediate activation of macrophages in response to infection, their associated susceptibilities to human immunodeficiency, hepatitis and West Nile viruses suggesting they impact virus-related aspects of the immune-inflammatory response (Aim-V or blue print in column H of S5 Table).

Genes more related to cellular autonomic-immune response (Aim) include *IFNAR1/2* or *STAT2* from above that regulate interferon action and *TNFRSF13B1* that participates in T-cell signaling, all three genes associated with immunodeficiencies in S5 Table. Other genes intimately related to viral response include the *ICAM1* and *IFITM3* genes encoding cell adhesion molecules involved in immunity, the interferon alpha-1 *IFNA1* gene associated with Epstein-Barr virus susceptibility, and the IRF9 binding component of the interferon-induced transcription factor complex that includes the *STAT2* gene from above (S5 Table). Many of these gene products like cell adhesion molecules would be targets for vaccines like the SARS-CoV-2 spike protein [58].

Many genes involved in COVID19 susceptibility have a similar breadth of associated diseases as those contributing to EDS, 13 having impact on embryonic development (-Dev in column F, S5 Table). These include the *LIFR* gene altered in both conditions (Fig 5) that is associated with multiple anomalies in Stuve-Wiedemann syndrome (M601559) and the *WNT3* gene associated with limb agenesis (M273395).

At least 26 of the COVID19 gene associations have symptoms of connective tissue dysplasia (red print in S5 Table column H) including particularly the *NLRP3* gene altered in both conditions (Fig 5) that is associated with joint pain, muscle aches, and symptoms of mast cell activation (M191900); note also the *ATP6V1A* gene that is associated with a lax skin disease (M617403) with tall stature, aortic dilation, and joint contractures. Another 7 associations in S5 Table involve molecular similarities with EDS-relevant disorders (red print underlined), the *FURIN* (M136950) and *TEAD3* (M603170) genes impacting transforming growth factor-beta pathways (as do the *FBN1* and *TGFB/R* genes altered in EDS patients), the *DPP7* (M610537) and *DPP9* (M608258) peptidases that cleave proline residues abundant in collagens [128], the *DDR1* receptor (M600408) that binds fibrillar collagens [15], the lung surfactant protein *SFTPD* (M178635) that has collagen-like glycine-hydroxyproline-hydroxylysine residues, and the *NDUFAF79* gene (M615898) involved in assembly of mitochondrial complex I--recall Fig 3 showing 31 EDS patients with *MT-ND* component gene alterations.

Especially of interest based on the renal complications of COVID19 are 8 genes associated with renal disease (purple print in S5 Table), 4 of them impacting vessels that include *ACE1*, *AGT*, *AGTR1* related to angiotensinogen-angiotensin I/II conversions and the SARS-CoV-2 nasal epithelial receptor gene *ACE2* that is homologous to *ACE1*. The latter product is a metalloproteinase that is also expressed in the vascular endothelium of heart and kidney; it has not yet been associated with a hereditary disease despite its location on the X chromosome. The *ADAMTS13* gene similar to *ADAMTS2* in Fig 5 is associated with a clotting diathesis and renal disease (M274150) that could relate to thrombotic complications of COVID19 [53]—see below. Three genes in EDS patients affect the kidney (S2 Table), the sodium-chloride co-transporter *SLC12A3* associated with Gitelman syndrome M263800, uromodulin *UMOD* associated with renal tubular disease M263800, and *PKD1* associated with polycystic kidney disease M173100 (classified as having vascular impact like the COVID19-associated genes above).

Ten genes are associated with neurologic disorders (green print in S5 Table), including the *IRF3* gene associated with an encephalopathy (M616532) conferring headaches and brain fog and the *RAB7A* gene with a form of Charcot-Marie-Tooth disease (M600882) that is familiar from EDS discussion. The apolipoprotein E protein is associated with Alzheimer (M104310+) and heart disease (M617347) and the NPC1 cholesterol trafficking regulator that allows lysosomal accumulation in Niemann-Pick disease (M257250). These have similar actions to the *LPIN1* (M605518) and *LDR* (M606945) genes variant in EDS patients (S2 Table).

The X chromosome androgen receptor AR that like many EDS-contributing genes impacts muscle (M313200+), joins *ACE2*, *TLR7* and 6 other X chromosome genes as potential factors in male susceptibility to COVID19. The latter contrasts with the 85% female preponderance in EDS (Table 1) although sex ratios in long COVID19 may be more equal (see below).

Fig 4A indicates that 21 of the genes influencing COVID19 infection are judged more related to viral entry/proliferation (virulence) and 83 more related to host responses that could mimic mechanisms of EDS-dysautonomia. Even with this exclusion, Fig 4A shows that genes related to immune-inflammatory processes (Aim) by their prior disease associations comprise 26 of 83 or 31% of those influencing COVID19 in S4 Table versus 8.8% of the 317 contributing to EDS in S2 Table. Proportions of genes impacting other elements or processes in Fig 4A are similar for cardiovascular (42 or 13% versus 10 or 13%), neural (101 or 32% versus 26 or 31%), clotting (4.1 versus 4.8%), and skin (3.5 versus 4.8%). They differ significantly (red or green circles) in categories of other autonomic (14 versus 2.4%) or muscle (11 versus 3.2%) and substantially for bone (22 versus 1,2%), joints (6.3 versus 2.4%) and renal (0.95 versus 4.8%).

Gene product types may reflect the importance of structural proteins in EDS and immune signaling after COVID19 infection, 76 or 25% of EDS-relevant genes being structural (St) versus 6 or 7.2% for COVID-relevant, 39 or 12% of the former having signal (Si) functions versus 20 or 35% of the latter in Fig 4B. Other product proportions are similar, including the 3 (11%) of 82 COVID19-related genes and 40 (13%) of 317 EDS-related genes with mitochondrial connections: *STAT2* (elongated mitochondria in muscle), *TLR3* (mitochondrial antiviral pathway), and *NDUFAF7* (assembly of mitochondrial complex I) are related to COVID19 infection (S5 Table) and similar to EDS-relevant genes (Fig 5).

### Comparing individual symptoms and genes relevant to COVID19 with those of EDS

As stated in the Introduction, a pattern of persisting symptoms dominated by fatigue, brain fog, breathing problems, and joint-muscle pain became apparent in patients recovering from SARS-CoV-2 [37–42]. This finding constellation became known as post-acute COVID19 sequalae (PACS) or long COVID19, a recent report estimating that 6.2% of people had one of three symptom clusters (persistent fatigue with bodily pain or mood swings, cognitive problems, ongoing respiratory problems) after COVID19 infection [43].

The variable periods, vacillating intensity, and subjective nature of long COVID19 symptoms have been difficult to characterize, but a unifying theme is autonomic dysfunction as demonstrated by measures of orthostatic intolerance and postural orthostatic tachycardia syndrome [38–42]. The systematic review of Deer et al. from 2021 [37] adopted standard phenotypic descriptions for symptoms [129] and included 59 articles among 303 that looked at clinical manifestations 3 weeks or more after initial symptoms of COVID19 infection (outpatients) or hospital discharge (inpatients).

As with the EDS patients described here, the 81 cohorts reviewed [37] were heterogenous with mixes of post-infectious timing, outpatient-hospital-intensive care, physical examination-self report, sex, and age (overall male to female ratio of 1.2 to 1 estimated from their data). Also similar were variable frequencies of laboratory-pathology findings--some suggestive of long-term organ damage after COVID19--and the inevitable ambiguity of symptom descriptions [fatigue-how chronic, steatosis or fatty liver?—37]. Although standard nomenclature for symptoms [129] is an asset, it does not group symptoms by clinical mechanism.

### Comparison of EDS and long COVID19 symptoms

Symptoms common to EDS and long COVID are shown at the top of Fig 5 using the analogy to Tolkien’s Ents: A tissue laxity-dysautonomia entome is imagined with a converging network of contributing genes at the bottom (roots) and a diverging network of symptoms at the top (branching canopy), the two connected through major pathophysiologic mechanisms like articulo-autonomic dysplasia (flowing channels of phloem or sap in the trunk). Peripheral genes with less impact on the central mechanism will have less disruptive variations in affected patients while those like *COL3A1* (M120180) will act as nodes in these gene networks and cause more numerous and severe symptoms.

The large percentage ranges for symptoms in both patient groups reflects the heterogeneity of patient ascertainment (clinic, online, retrospective in EDS, different hospitalized-outpatient cohorts, post-infection times for COVID19), and the subjective nature of reported findings. All symptoms, ordered by percentage in COVID19 patients, are more frequent in EDS although ranges are a bit more compatible. Symptoms of autonomic imbalance (brain fog, chronic fatigue, asthma-dyspnea, sleep difficulties, and tachycardia) are common in both EDS and long COVID19 (Fig 5), compatible with a prior hypothesis [42]. Asthma is a consequence of mast-cell activation [32–33], the other four of postural orthostatic tachycardia syndrome [30–31, 39].

Less common in long COVID19 than EDS are IBS symptoms and those orthostatic hypotension like syncope and dizziness. Neurologic symptoms like difficulty walking-poor balance, muscle weakness, myalgia, and frequent headaches occur in both as does joint pain that is common in EDS, post-infectious, or autoimmune illnesses (Fig 5). Occurring occasionally but not chronic in EDS are the cough (16%), chest pain (14%), congestion (10%), sore throat (4%), and low-grade fever (4%) reported by Deer et al. [37], symptoms possibly related to persisting viral infection.

### Similar genes relevant to EDS and COVID19 severity

Genes highlighted by variance or expression in both disorders include the *F2* prothrombin gene (M176930) related to bleeding disorders and the metalloproteases *ADAMTS2* (M6045539) and *ADAMTS13* (M604134), the latter gene product interacting with the von Willebrand factor that had 18 coding variants in EDS (Table S2). Ratios of the ADAMTS13 and VWF proteins are related to thrombosis and COVID19 mortality [52–53], recalling the 13 genes and 34 variants in EDS patients that impact clotting functions (S2 Table, the 15 patients with *VWF* gene variants in Fig 3).

The shared *LIFR* leukemia inhibitory factor receptor (M151443) with immunoglobulin/cytokine domains and the *NLRP3* (M606416) pyrin-like genes could be involved in the inflammatory response to COVID19 as well as the enhanced inflammation from adrenergic stimulation in EDS and other conditions [10-11, 23, 77]. Similar dualities for the *STAT1* (M600555), *TNFRSF13B* (M6049097), and *TICAM1* (M607601) genes may apply since the first two are associated with immunodeficiency disorders and the last confers susceptibility to encephalopathy from herpes virus infection (S5 Table).

Among the 18 similar genes are complement components *C3* (M120700) and *C1R* (613785) that with the *NFKB1* (M164011)-*NFKB2* (M164012), *IFR3* (M603734)-*IF1H1* (M606951) genes (Fig 5) and others could also mediate inflammatory and autoimmune symptoms. The *POLG* (M174763) and *NDUFA11* (M612638) genes variant in EDS are most easily related to neurologic and autonomic symptoms, the similar *POLD4* (M611525) gene definitely (S5 Table) and the *NDUFAF7* (M615898) likely having neurologic impact [50]. The *PIK3C3* (M602609) gene similarity to *PIK3R1* (M171833) that is associated tissue laxity is an example of 28 COVID19-related genes having molecular or symptom similarities to connective tissue dysplasias (red print in S5 Table).

## Discussion

This clinical genetic study of EDS relates its quantified finding pattern to underlying articulo-autonomic dysplasia mechanisms and the multiple gene variants found by NextGen DNA sequencing. Major results are the connected tissue laxity-neural symptoms of EDS, their relation to disparate nuclear and mitochondrial genes, and the similarities of these relationships to those of acute or long COVID19. The study illustrates the potential strengths and limitations of genomic analysis as summarized below.

### Clinical-DNA correlation in EDS

This uniquely large study of 1899 EDS patients, 1261 by systematic evaluation, is still limited by heterogenous settings (outpatient versus online in Table 1) and subjective symptoms (S1 Table) that set the stage for standardized prospective studies [130, 131]. Strengths include the holistic ascertainment of syndrome pattern, patients referred by self, general, or subspecialty physician having as many neuromuscular (98%) and dysautonomia (96%) findings as those traditionally emphasized in the joints (99%) or skin (93%, S1 Table).

The large standard deviations for numbers of symptoms in various history and physical categories (Table 1) compromise group comparisons but indicate significant differences between women, men, those under 10.5 years, and those not meeting EDS criteria [66]. Exclusion of integral neuromuscular findings likely accounts for the 7 to 13-year diagnostic delays of EDS patients, their frequent dismissal by physicians leading to exceptional gratitude for diagnosis. Dysautonomia findings like brain fog (64-83%), chronic fatigue (64-87%), or bowel irregularity (75-82%) are as common as hypermobility (60% > Beighton 7) in EDS patients (S1 Table) and must be part of their medical evaluations or future studies.

Complete ascertainment of patient findings and their relation to pathogenetic mechanisms is necessary for understanding genetic influence on diseases like EDS as shown by the molecular *<u>and</u>* medical protocol of Fig 1. This clinical qualification of DNA variants, added to consensus qualifications [83–84, 92] is intended to overcome medical distrust of genomic results [94] by 1) minimizing use of the unhelpful variant of uncertain significance qualification, 2) adding connotations for less helpful variants (VUDU, VnoDU) or those suggestive of dual diagnoses (V*DUO), and 3) emphasizing that DNA changes may support but never make a “molecular” diagnosis [92] without experienced clinical correlation.

DNA variants become candidates for disease correlation in the way their genes used to become candidates for products or loci before genomics. Correlation with pathogenic mechanism is emphasized more than functional analysis, the former difficult to model *in vitro*, the latter reserved for research that will ultimately “elect” or reject candidate gene relevance to a particular disease. Even patients with homozygous mutations associated with sickle cell anemia may have minimal symptoms of that disease [95], their contribution to diagnosis needing additional clinical judgment.

Underlying articulo-autonomic dysplasia mechanisms related 317 variant genes to EDS (S2 Table, Fig 2), additional studies needed to validate contribution of 175 genes with single variants in that Table. The 26 EDS-related genes encoding transcription factors in Fig 4 predict discovery of additional regulatory region variants, an explanation for the 41% EDS patients not having variants found by whole exome sequencing. Although shared genes and different sex ratios compromise the comparison, relevance to tissue laxity-dysautonomia symptoms of 893 relevant DNA variants found in 568 EDS patients is supported by their differences from 154 DNA sequence variants found in 82 disability patients (Tables 2-3, S2-S4).

Recurring variants in Fig. 3, S2 and S3 Tables also support EDS association, from the 15, 13, and 51 patients with variants in the *COL1*, *3,* and *5* genes previously associated with EDS [5, 7, 98] to patients with newly associated variants in the *COL7*-*17*/*FLG* (40-skin/inflammation), *SCN5*/*9*/*10*/*11*A (24-nerve), *MT-trRNA*-*CO-ND-CYB*/*COL6*-*12* (153-muscle), *COL9* (8-bone), *VWF*/*FBN1*/*TGFB* (49 clot-vessel), and *MT-ATP6*/*POLG*/porphyria (67-autonomic) genes that suggest action through a tissue element/process network.

Another advantage of this pattern-mechanism approach is its unifying qualification of previous (*FLNA*, *ATP7A*) or current (*ADAMTS2*, *FKBP14, PLOD1*, *TNXB*) type-associated variants with more common and congruent EDS profiles. Participation in a tissue laxity-dysautonomia gene network is also attributed to the *ABCC6*, *COL1*, *COL3, FBN1*, and *TGFB* genes usually associated with pseudoxanthoma elasticum [103], osteogenesis imperfecta [98–99], vascular EDS [7, 100], Marfan [80], and Loeys-Dietz [107] syndromes by their variants in S2 and S3 Tables. Network action implies impact of heterozygous variants, even those previously associated with recessive disease [104]. as supported by the similar finding profiles of Fig. 3 that need larger patient numbers for significance.

### Distribution and nature of EDS and COVID19-related genes

The genes associated with EDS (317) or COVID19 severity (104) are distributed on all chromosomes (excepting Y and 8, 13, 16, 20 for COVID19) with clusters at 2q32.2 (*COL5A2/COL3*), 3p24.1 (*SCN5/10/11A*), 11q23.3 (*SCN2/4B*), and 21q22.3 (*COL6A1/A2*) for EDS and at 3p22.2 (*CCR1/5-CXCR6*) for COVID19 (Fig 2A). Genes impacting mitochondrial function include 30 of 37 in mitochondrial DNA (Fig 2B), 10 EDS-related (*NDUFA*-*11*/*S3*, *OPA1*, *TYMP*, etc. green print), and 3 COVID19-related (*STAT2, TLR3, NDUFAF7*) in nuclear DNA Fig 2A. The encoding of products with structural (*SURF1*, *MT-trRNA*), respiratory enzyme component (*MT-ND*/*CO*), adhesive (*NUBPL*), or DNA polymerase (*POLG*) functions by these genes suggests influence on EDS by depletion of mitochondrial number and/or energy coupling. Mitochondrial roles in aging [118] and immunity-inflammation [119, 124–125] may explain influence on COVID19 infection.

Diversity of function (S2, S5 Tables, Fig 4) and location (Fig 2A) of genes influencing EDS or COVID19 is consistent with their participation in networks regulating connective tissue integrity and its reciprocal autonomic regulation. Both functions would be impacted by gene variation in EDS while autonomic imbalance with its immune and inflammatory dysregulation would be more impacted in COVID19. A primordial regulatory-structural operon might be imagined for initial metazoan transitions [14], duplication and realignment of protein domains shown by the binding of acetylcholinesterase to collagen by COLQ protein [22], the interspersion of VWF motifs in COL3 [17] and COL7 [20] proteins, the service of abundant collagen type I as anchor for immune molecules [15] and core for other types during fibril formation [97]. This modular pleiotropy is supported by the 184 (62%) of 298 associated disorders with at least 3 tissue dysplasia symptoms in S2 Table (orange shading).

Attribution of variant genes in EDS to tissue element or process (Fig 4A) fostered comparison to the 104 relevant to COVID19 severity (S5 Table), 18 genes similar and 6 identical between the two groups (Fig 5). These include variant genes with parallel impacts or functions like *ADAMTS2/F2/PIK3R1* influencing EDS and *ADAMTS13/F2/PIK3C3* influencing COVID19 that impact clotting-tissue laxity, *LIFR/NLRP3/STAT1/T1CAM1/TNFRSF13B* (both) plus *C1R/IF1H1/NFKB2* (EDS) and *C3/IFR3/NFKB1* (COVID19) that impact immunity-inflammation, *SLC6A2* (EDS) and *SLC6A20* (COVID19) that have transport functions, and *POLG/FOXP2/RBM20/WNT10A/ZNF469* (EDS) and *POLD4/FOXP4/RBM15/WNT3/ZNF275* (COVID19) that have DNA polymerase/regulatory functions (Fig. 5, S2 and S5 Tables). The occurrence of small fiber neuropathy [12, 56] and thyroid dysfunction [S1 Table, 116-117] in EDS (Table S1) and COVID19 [59] along with the many shared joint-muscle and dysautonomia symptoms in Fig 5 support the operation of overlapping gene networks in these disorders.

### Connected findings and genes as “entomes”

These networks may be analogized to Tolkien’s Ents, their genes as rhizomes, clinical mechanisms as trunks, medical problems as the diverging branches of the entome (Fig 5). Clinical findings caused by these overlapping gene networks will have the opposite branching, widely shared traits like whole body pain, muscle weakness, and adrenaline surges (stems) being more frequent in EDS or long COVID 19 than their component symptoms of arthralgia, myalgia, headaches, poor balance, or chronic fatigue (leaves, upper part of Fig 5).

Entomes differ from gene modules or molecular pathways by connecting genes to sign and symptom patterns. Genes converge to and symptoms diverge from central pathogenic mechanisms, key genes and common symptoms being nodes of their respective networks (lower part of Fig 5). The idea of entome connects these mirroring networks of genes and symptoms through pathogenetic mechanism, their divergent clinical findings like the distributed flood debris that can be related to normal structures only by knowledge of floodwater force and direction.

Analogies to Ent motion involve centuries of gene (rhizome) evolution at one extreme and the daily changes in physiology (trunk-phloem) and symptoms (canopy) at the other. Networks affecting connective tissue likely arose in early metazoan/metameric evolution [14] and were tailored to produce the later upright posture, joint mobility-dexterity, and forward-facing binocular vision of primates [132, 133]. Necessary balance between the tissue-orthostatic stability enabling forward visual accommodation and the flexibility needed for ambulation and limb reach/grasp is shown by the articulo-autonomic dysplasia symptoms of S1 Table.

### Implications for future research

#### Expanded studies of EDS and long COVID19 symptoms and outcomes

Systematic ascertainment of patient findings via the findings of S1 Table included few laboratory and no medical measures like the results of echocardiography-vascular screening [7], tilt-table [30, 31], nerve conduction [13], intestinal motility [36], or imaging for Chiari [134], median arcuate ligament [135], or nutcracker changes [136] that lead to immediate therapies. Evaluation protocols adding the latter results could improve symptom descriptions [129] and outcome measures [130] for EDS and long COVID19 as done for COVID19 infection in patients with rheumatic diseases [131]. Better documentation of severe complications like aneurysms or cardiomyopathy that were incidentally mentioned in 23 or 2 of 1261 EDS patients (S2 Table or data not shown) would better discriminate the patients with *COL3* mutations in Fig 3 from those with the vascular type [7, 100]. When these more complete protocols were used for patients with EDS and COVID19 infection, their objective clinical profiles could be compared to those of other infectious conditions like multisystem inflammatory syndrome in children [137] and Kawasaki disease [138]. COVID19 hospitalization and mortality were not increased in patients with fibromyalgia [55], but this symptomatic and heterogenous diagnosis ignores many findings of EDS-dysautonomia and has limited association with biomarkers [139].

#### Expanded and enhanced DNA databases

This large patient collection barely sketches the genomics of EDS and shows the massive numbers of appropriately qualified DNA testing results that will be needed to provide understanding, diagnosis, and informed management of multifactorial disease. The need for clinical correlation of DNA variants is underlined by several genetic properties reviewed in this study. Not only are different connective tissue dysplasia phenotypes produced by different types or locations of mutations in the same gene [24-27, 35, 98, 103, 105, 108-111, 115, 123], but many component-fabricated genes like collagens [17,20] will have shared domains that could be mutated to give similar phenotypes. These may also result from mutations in different collagen genes since several types of collagens participate in fibril assembly [18, 19, 97]. These considerations make one gene-one type/disease matches [6–7] unlikely and molecular diagnoses without clinical correlation [92] untenable for EDS and most likely for any genetically influenced disease. Collaborative interpretations of variant diagnostic utility-disease relevance by molecular geneticists *<u>and</u>* appropriate physician subspecialists per the Fig 1 protocol are required if DNA testing is to become a prime contributor to precision medicine.

#### Explore mitochondrial and neuromuscular function in EDS and COVID19

The 135 EDS-related genes impacting neuromuscular elements, the 71 associated with autonomic imbalance, and the 40 influencing mitochondrial function (Fig 4, S2 Table) emphasize decreased muscle constraint of connective tissue and its vasculature as a key cause of joint-skeletal and dysautonomia symptoms. Many genes like *FLNC* in Table 4 are associated with cardiomyopathy and muscle weakness, reflecting overlap of proteins in cardiac, skeletal, and probably intestinal smooth muscle (possibly contributing to the 92% of EDs patients with bowel dysmotility in S1 Table). Further study of musculoskeletal and mitochondrial dysfunction [140–141] in EDS and acute/long COVID19 could justify trials of promising dietary [31], physical therapy [18, 142] and exercise [75, 121, 143] protocols in both disorders. Important objectives regarding long COVID19 are to associate symptom frequencies and outcome measures with defined post-infection time periods, then determine whether the genes influencing COVID19 infectious (S5 Table) also influence the duration and disability of its post-infectious phases.

#### Future therapies for EDS, COVID19, and the related symptoms of aging

Given the similarity of many EDS symptoms like skin laxity or poor balance to those in the old [11] and the elder vulnerability to COVID19 [44, 54], will the 108 genes that impact connective tissue elements in Fig. 4 (EDS), the 28 associated with connective tissue dysfunction in S4 Table (COVID19), and the implications of mitochondrial dysfunction [118]in both disorders indicate kindred mechanisms in aging? If so, then unified therapy approaches could be applied to the flexible [4], frail [54, 121], or infected [39] that could, as basic science distills cause from the present correlations, involve autologous transplants with variant-edited [144] mesenchymal stem cells [29, 145].

## Data Availability

Sharing of the EDS1261GW1-23 database with deidentified clinical information is offered to those emailing the author. ClinVar and Mitomap database numbers are given for those DNA variants that could be found (∼70%). The entire DNA variant files for EDS patients will be offered to these databases with clinical relation to Ehlers-Danlos syndrome when vaidated by accepted for publication by this or another journal. All relevant data are within the manuscript and its Supporting Information files.

## Supporting information

Supporting information includes an Appendix (Supporting information Appendix for a shared EDS-COVID19 gene network.docx) containing 5175 words and an Excel file (Supporting information S1-S5 Tabs for EDS-COVID19 gene network.xls) that contains 5 supplemental S1-S5 Tables and, for review, Sheets 6 containing the EDS1261GW database with deidentified patient numbers, age ranges, sex, findings, and positive-negative DNA results without gene-variant details. This is the database that will be mailed to scholars by request to the author along with the key to encoded findings placed in Sheet 7 of the Excel file for review. The Supporting Information tables meant to be published if accepted are:

**S1 Table. History and physical finding frequencies in 1064 EDS females and 197 EDS males with systematic evaluations**

**S2 Table. Genes variant in EDS with their descriptions and associated diseases**

**S3 Table. Primary and additional DNA variants found in EDS patients listed by patient number**

**S4 Table. DNA variants in patients with developmental disability**

**S5 Table. Genes relevant to COVID19 infection severity (with references to specialized articles below the table)**

## Acknowledgements

Collaboration of Dr. Vijay S Tonk on the protocol for variant qualification, assistance of genetic counselor Melissa Alderdice and others at the GeneDx Company© with coordination of genetic testing, referrals and insights from cardiologists Amer Suleman and Lee Ann Pearse, orthopedist W. Barry Humeniuk are appreciated.

